# SARS-CoV-2 Suppression and Early Closure of Bars and Restaurants : A Longitudinal Natural Experiment

**DOI:** 10.1101/2021.08.07.21261741

**Authors:** Reo Takaku, Izumi Yokoyama, Takahiro Tabuchi, Masaki Oguni, Takeo Fujiwara

## Abstract

Despite severe economic damage, full-service restaurants and bars have been closed in hopes of suppressing the spread of SARS-CoV-2 worldwide. This paper explores whether the early closure of restaurants and bars in February 2021 reduced symptoms of SARS-CoV-2 in Japan. Using a large-scale nationally representative longitudinal survey, we found that the early closure of restaurants and bars decreased the utilization rate among young persons (OR 0.688; CI95 0.515-0.918) and those who visited these places before the pandemic (OR 0.754; CI95 0.594-0.957). However, symptoms of SARS-CoV-2 did not decrease in these active and high-risk subpopulations. Among the more inactive and low-risk subpopulations, such as elderly persons, no discernible impacts are observed in both the utilization of restaurants and bars and the symptoms of SARS-CoV-2. These results suggest that the early closure of restaurants and bars without any other concurrent measures does not contribute to the suppression of SARS-CoV-2.

## 1 Introduction

During the fight against severe acute respiratory syndrome coronavirus 2 (SARS-CoV-2), almost all developed countries shut down full service restaurants and bars, or implemented strong restrictions on their functioning(Madeira, Palrão and Mendes, 2021; Yang, Liu and Chen, 2020; Fairlie, 2020). Even if they were allowed to be open, their operating hours were restricted. Because the restaurant industry occupies a large share of employment, these policies would increase the unemployment rate (Kong & Prinz, 2020) and firm exit (Miyakawa, Oikawa and Ueda, 2021), and lead to the deterioration of the mental health of workers in this industry (Witteveen & Velthorst, 2020). In Japan, even with the high rate of vaccination, full service restaurants and bars were under strict restrictions from July 12 to September 30 2021 due to the spread of the Delta variant. On January 2022, several prefectural governments including Tokyo began to implement strict measures for the restaurant industry again due to the spread of the Omicron variant.

Despite the significant economic damage, these policies were implemented in hopes of suppressing SARS-CoV-2 because plenty of scientific research suggest that full service restaurants and bars contributed to the spread of SARS-CoV-2 (Chang et al., 2020; Fisher et al., 2020; Brauner et al., 2021; Askitas, Tatsiramos and Verheyden, 2021; Haug et al., 2020; Persson, Parie and Feuerriegel, 2021). However, it is not clear whether this early evidence can be directly applied to the situation today for several reasons. First, these studies are based on data from the first wave when many other non-pharmaceutical interventions (NPIs) were jointly implemented (Brauner et al., 2021; Hale et al., 2021). Thus, it is quite difficult to separate the effects of closing restaurants and bars from the effects of other concurrent policies. In addition, during the first wave, many people in North American and European countries did not notice the importance of wearing masks (Howard et al., 2021; Kwon et al., 2021; Cheng et al., 2021) and washing their hands frequently (Makhni et al., 2021), which was proven to be effective in preventing SARS-CoV-2 afterwards. This situation is also quite different from that in the second or later waves, when most people voluntarily performed several preventive behaviors that were effective. Therefore, it is of particular importance to examine the effects of strict restrictions on restaurants and bars with much more timely data and better experimental settings.

During January and February 2021, Japan had a unique opportunity to serve as an ideal experimental laboratory to investigate the effects of strong restrictions on restaurants and bars. During this period, there were neither strict NPIs, except for the early closure of restaurants and bars, nor city lockdowns, border restrictions, school closures, and stay-at-home orders with financial penalties. Japan’s negligence toward countering the spread of SARS-CoV-2 contrasted from that of other developed countries (Hale et al., 2021).

Instead, in Japan, 11 prefectures in Figure 1 declared a state of emergency (SE) in January and implemented strict restrictions on full service restaurants and bars. Specifically, 11 prefectural governments prohibited restaurants and bars to serve alcoholic beverages after 7 p.m., and forced them to close by 8 p.m., compensating for their financial loss with generous governmental subsidies. Among the 11 prefectures, the SE was lifted off only in the Tochigi Prefecture on February 7th, while other prefectures continued till the end of February. The local SE administration in Japan only asked the general population to stay at home voluntarily, without any explicit legal enforcement (Hosono, 2021; Watanabe & Yabu, 2021).

**Figure 1:**
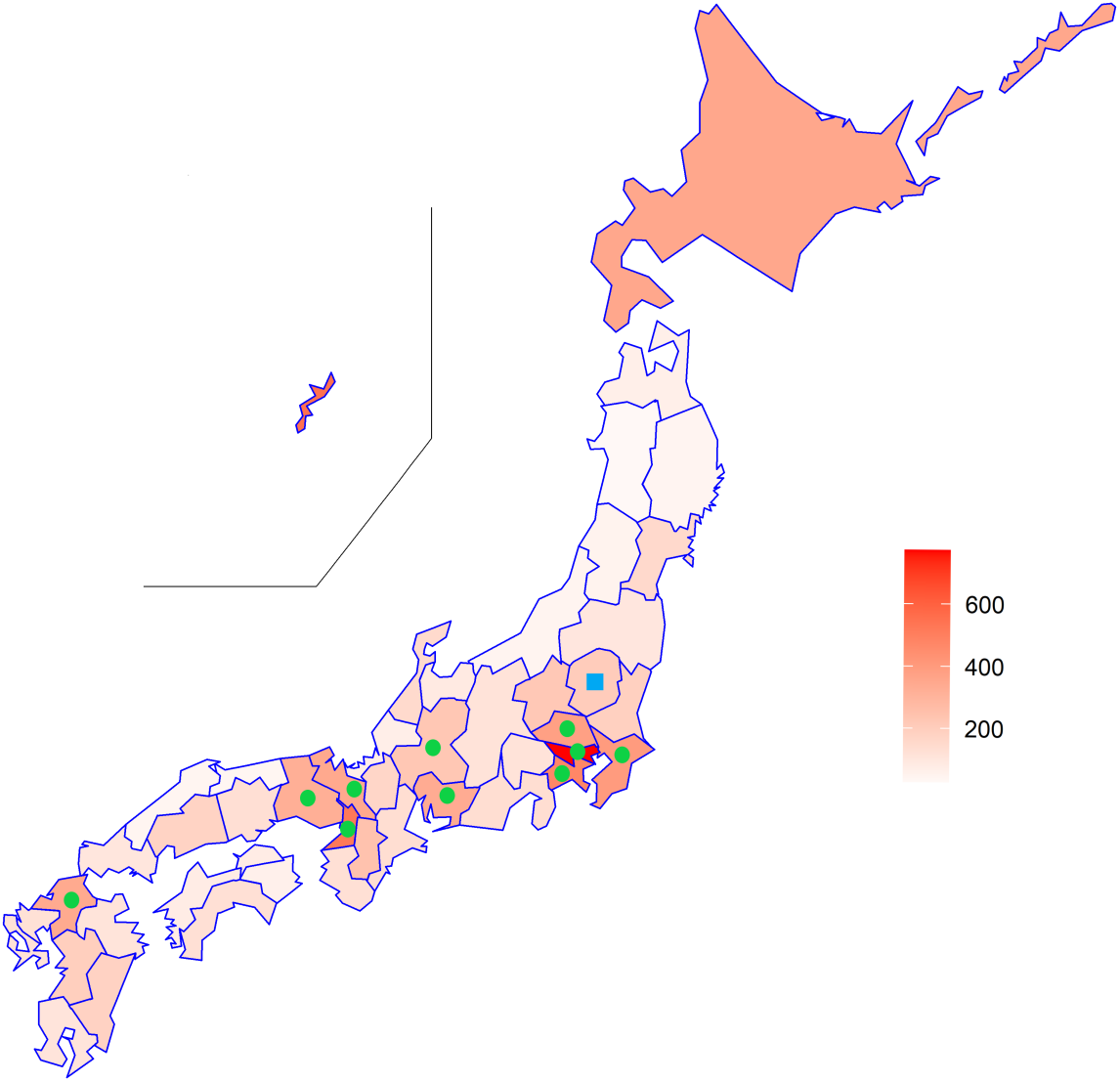
Infection Rate across Prefectures. *Note;* The infection rate is per 100,000 persons. Circles represent the 10 prefectures that declared SE in January and continued till the end of February. The square represents Tochigi prefecture, which declared the SE in January but lifted it on February 7th.

How does this policy of targeting restaurants and bars affect virus transmission? The answer hinges on how the people affected by this policy, namely those who frequently visited restaurants and bars, altered their behavior. One scenario is that they had to stay at home and not go anywhere at night. In this scenario, the targeting policy effectively suppresses SARS-CoV-2 transmission. However, this is an optimistic view of the effectiveness of the policy. In reality, the people who were affected by this policy were not scared of SARS-CoV-2 generally, and behaved carelessly and spread the infection. For example, many people celebrate the new year with relatives and friends in January, and it is possible that new year parties were held in changed venues rather than being canceled altogether. In this case, the effectiveness of SARS-CoV-2 suppression would be quite limited.

In addition, it should be noted that, unlike the first wave, many individuals started with a much higher awareness of preventive behaviors in the third wave. Thus, it should be re-examined whether the early closure of restaurants and bars, in addition to a high standard of preventive behavior, reduced the spread of SARS-CoV-2 at the same level as predicted in the first wave. Consistent with this view, evidence from the UK found that early closure of restaurants and bars in September 2020 had no measurable effect on person-to-person contacts (Jarvis et al., 2021), because the level of contacts was already quite low after the first wave. Even in the evidence from the first wave, some studies find that the effects of closing restaurants and bars are limited when other strict measures are implemented (Post et al., 2021).

Most importantly, previous studies consistently pointed out that people reduced their mobility mainly because of the fear of SARS-CoV-2, rather than governmental interventions such as shelter-in-place orders (Goolsbee & Syverson, 2021; Berry et al., 2021; Sheridan et al., 2020). Therefore, policies targeting restaurants and bars would have a very small impact on overall mobility and person-to-person contacts, without a strong fear of SARS-CoV-2 itself. To investigate the impacts of this policy, we estimated the difference in subjective symptoms of SARS-CoV-2 (Struyf & Van den Bruel, 2021; Miyawaki et al., 2021) between individuals under the Japanese SE declaration and others. Given that many studies suggest that the fear of SARS-CoV-2, not policies, induced voluntary reduction of mobility (Goolsbee & Syverson, 2021; Yan et al., 2021), a naive comparison between persons within prefectures with SE declaration and those without may contaminate the effects of the policy with the direct effects of fear itself. In fact, the early closure of restaurants and bars was implemented because of the spread of SARS-CoV-2; thus, it is still challenging to separate the effects of the policy from the direct effects of the virus, even if no other strict policies were implemented.

To overcome this challenge, we focus on individuals who live close to the borders of the prefectures with SE declaration and those without, by using the zip-code of respondents, in the same spirit as the geographical regression discontinuity analysis (Dell, 2010; Dell, Lane and Querubin, 2018). This empirical strategy seems to be useful because the fear of infection would be sufficiently similar in geographically proximal areas. Another feature of our methodology is that we use nationally representative longitudinal data with a short interval of only four months. Because the interval between the first and second waves is only four months, we could successfully absorb individual-level unobservables by incorporating individual fixed effects. With this difference-in-differences approach which focuses on geographically proximal areas, we study the effects of the early closure of full service restaurants and bars on (a) the utilization of full service restaurants (Japanese pubs) and bars in the past one month, and (b) 5 symptoms indicative of SARS-CoV-2, which are validated in previous studies (Struyf et al., 2020; Sonoda et al., 2021; Miyawaki et al., 2021).

## 2 Methods

### 2.1 Data

We study the effects of closing restaurants and bars by using a nationally representative internet survey, named the *Japan COVID-19 and Society Internet Survey (JACSIS)* (Zaitsu et al., 2021; Miyawaki et al., 2021). This web-based, self-reported questionnaire survey was administered by a large internet research agency Rakuten Insight, Inc.. The survey was conducted in accordance with the Declaration of Helsinki. Samples were coded and analysis was performed with anonymized database. Informed consent for survey participation was obtained from each patient. This study was approved by the Institutional Review Board of the Osaka International Cancer Institute (No. 20084).

The questionnaires were distributed to 224,389 individuals aged between 15-79 years old, and 28,000 responded during the first wave. The participants had the option of not responding to any part of the survey questionnaire, and discontinuing the survey at any point. The response rate was 12.5%. The surveys for the first and second waves were conducted in August and September 2020 and February 2021, respectively. Note that the second survey was conducted when the SE was declared in 10 prefectures, except Tochigi which declared the SE in January but lifted it in early February. From the dataset in the first wave, we chose 25,691 individuals, after excluding those with missing values. A total of 4,775 individuals were dropped in the second wave. The follow-up rate was 81%. Finally, we constructed a balanced panel data of 20,916 persons. The descriptive statistics are shown in SI Appendix Table SI1.

### 2.2 Main Outcomes

Our key dependent variable was the utilization of restaurants and bars within the past month. Specifically, restaurants and bars include *Izakaya* pubs, a common type of Japanese pubs that serve alcoholic beverages and a variety of tidbits. For simplicity, they will be referred to as “Japanese pubs” hereafter. Because the SE declaration in January and February 2021 explicitly prohibited these Japanese pubs and bars from serving alcoholic beverages after 7 p.m., and ordered them to be closed by 8 p.m., we expected the utilization rate to decrease. The 7 p.m. curfew for serving alcoholic beverages is crucial to the management of Japanese pubs, because Japanese people generally start drinking parties around 7 p.m. Therefore, curfews are regarded as a substantial business suspension order for Japanese pubs. In fact, a popular chain of Japanese pubs, *Watami*, completely shut down half their outlets.

On the health outcomes, we use 5 subjective symptoms indicative of SARS-CoV-2 (high fever, sore throat, cough, headache, and smell and taste disorder). Because there were a limited number of PCR tests in Japan, self-reported SARS-CoV-2 like symptoms have been regarded as a useful measure to detect persons with SARS-CoV-2 (Struyf et al., 2020; Nomura et al., 2020; Menni et al., 2020; Miyawaki et al., 2021). In addition, the validity of these symptoms has been confirmed in the environment in Japan (Sonoda et al., 2021). For example, for fever, the sensitivity was at least 80% in many studies, and the specificity was more than 90%. The specificity was also high for cough (at least 50%), sore throat (70%), headache (80%), and smell and taste disorder (90%) (Struyf et al., 2020; Sonoda et al., 2021; Miyawaki et al., 2021).

### 2.3 Research Design and Empirical Model

The intuition for this analysis is shown in Figure 2. In this figure, we plot the utilization rate of Japanese pubs and bars during the first and second waves, around the border of the 10 prefectures. On the horizontal axis, the negative (positive) sign represents the area without (with) the SE declaration. Because the SE was declared in urban areas, population density, which is shown in the histogram, increases as the distance to the border moves from -50 km to 50 km. Solid line represents the utilization rate in February 2021 (second wave) and dashed line represents the utilization rate in August-September 2020 (first wave). Reflecting an increase in urbanity, the utilization rate of Japanese pubs and bars exhibits an upward slope. Note that the utilization rate is about 36% at the border in both first and second waves, suggesting that the use of Japanese pubs is common in Japanese society. In addition, the utilization rate in February 2021, when the SE was declared, followed a parallel trend with that in August-September 2020, in the area outside the 10 prefectures. However, in the area inside the 10 prefectures, the utilization of Japanese pubs and bars drops to a larger extent, when compared to the trend in August -September 2020. In other words, the utilization rate of Japanese pubs and bars dropped only in the area under the SE declaration, when compared with the trend in August-September 2020. This pattern of the data strongly supports the view that the SE declaration has negative causal effects on the the utilization rate of Japanese pubs and bars, because within the short interval around the border, we can assume the the *fear* of infection (Goolsbee & Syverson, 2021; Yan et al., 2021) would be sufficiently identical.

**Figure 2:**
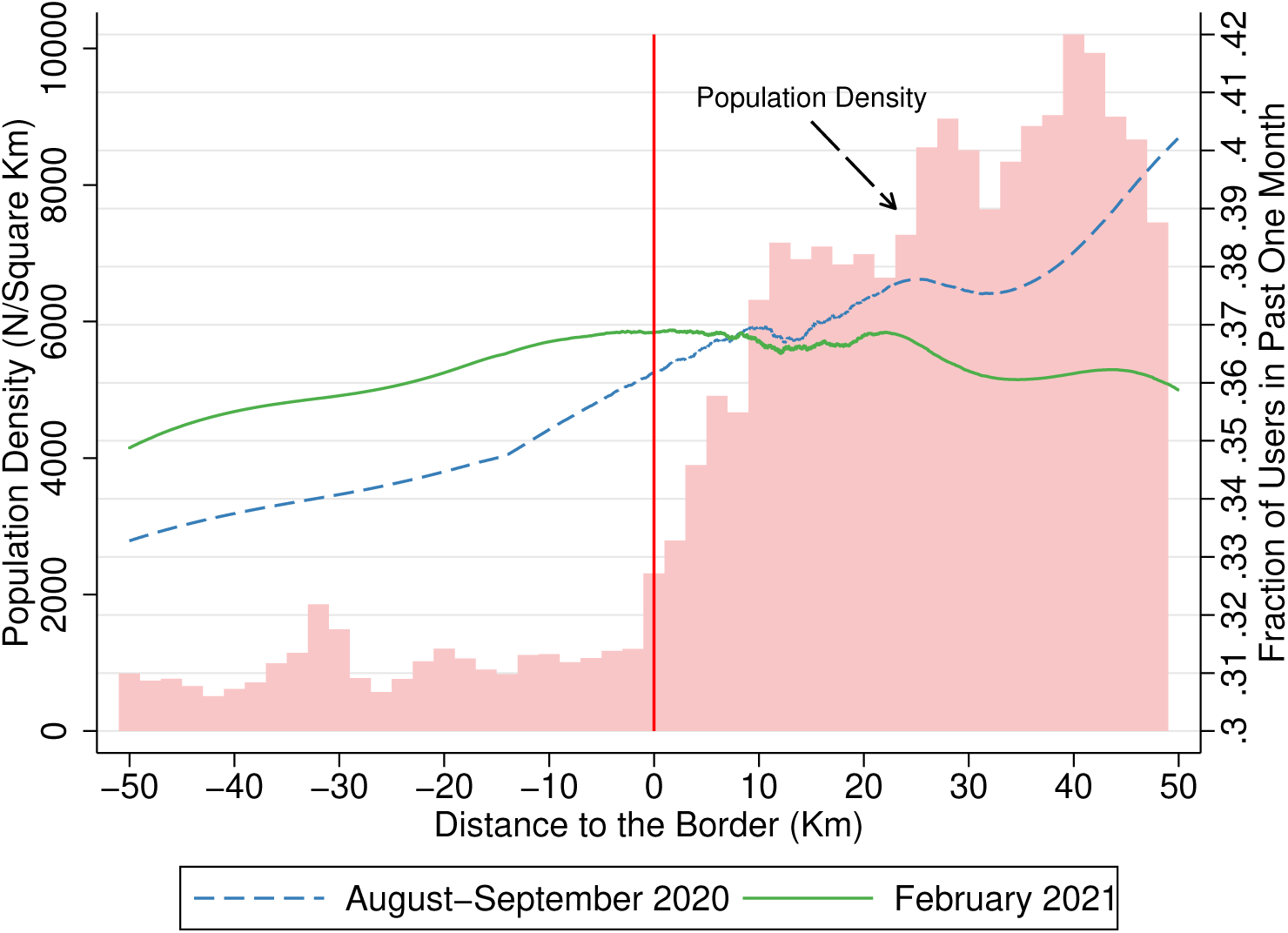
Population Density and Utilization of Japanese Pubs around the Border *Note;* The fraction of persons who went the Japanese pubs and bars are represented in lines which are derived from local polynomial fit. The vertical line represents the border that splits prefectures with (positive sign) and without (negative sign) the state of emergency declaration.

In the actual estimation to quantify the impacts of the SE declaration, we employ a difference-in-differences analysis with a fixed-effect linear probability model as well as a random effect logistic regression model, after adjusting individual level time varying covariates. In the case of a fixed-effect linear probability model, the DID specification is:

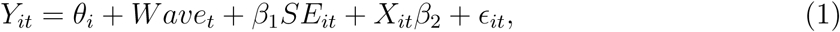

where *Y*_*it*_ is the outcome variable in individual *i* in wave *t*; *θ*_*i*_ is individual fixed effects; *Wave*_*i*_ is the wave fixed effects; *SE*_*it*_ is a binary variable which takes a value of one if individual *i* was under the SE declaration in wave *t*; *X*_*it*_ is a vector of individual level covariates. Specifically, we control for age, working status (i.e., job loss or not), marital status, smoking, and household income level. The household income level is measured by a binary variable that takes a value of 1 for households with an annual income of JPY 7 million (USD 70,000) or more. The descriptive statistics for these covariates are shown in SI Appendix Table SI1. Finally *ϵ*_*it*_ is an error term. In the actual implementation, we estimate equation (1) in the subsample of individuals who lived within the short interval around the border of SE declaration.

## 3 Results

### 3.1 Utilization of full service restaurants (Japanese pubs) and bars

We estimate the differences in the utilization of Japanese pubs and bars among persons who were differently exposed to the restrictive policy by using the fixed-effect linear probability model and random effect logistic regression model in Table 1. In Column (1), we find a statistically significant reduction in the utilization rate (OR 0.896; CI95 0.814 - 0.966), even with naive specifications, which compares the persons in the prefectures with and without the SE declaration.

**Table 1:**
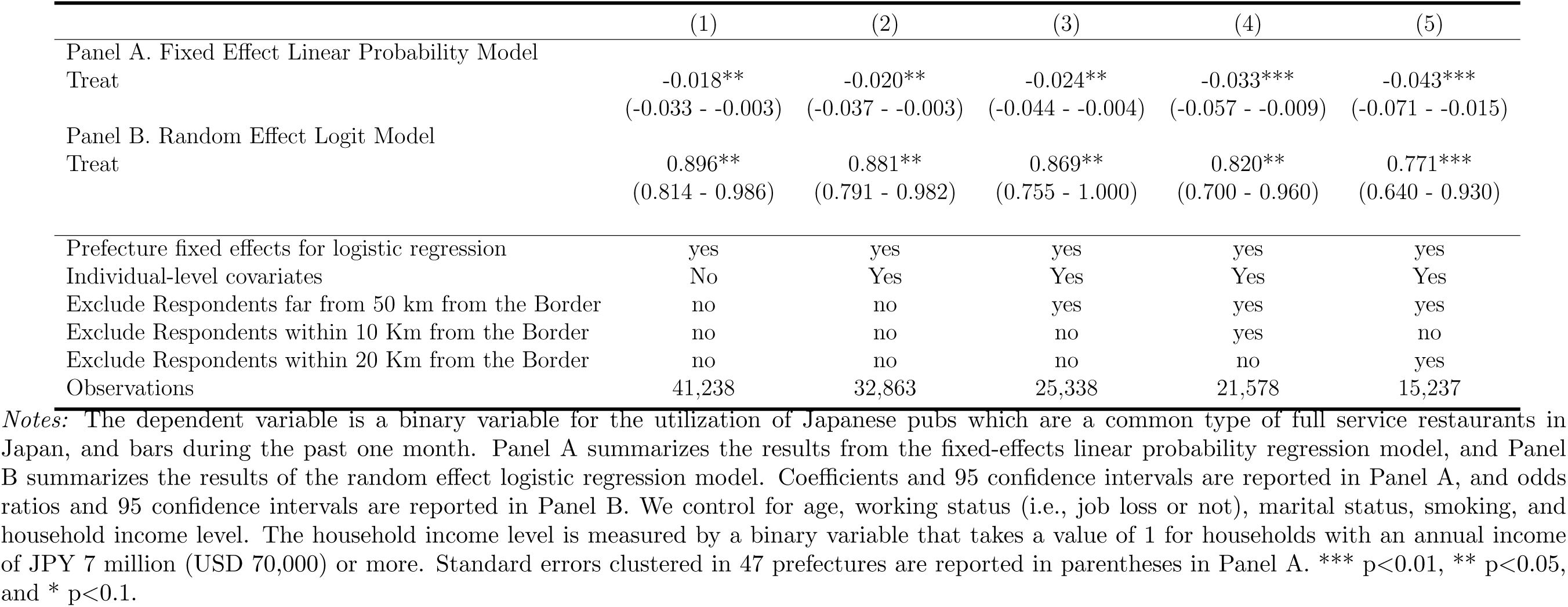
Effects on the utilization of Japanese pubs and bars

This result is robust when we additionally control for individual-level time-varying covariates in Column (2) and focus on the persons who live in within 50 km from the border, which separates prefectures with and without SE declarations in Column (3).

While the specification in Column (3) is robust for confounders such as direct behavioral effects from the virus itself (i.e., fear of the infection), we severely underestimate their impacts in Column (3) because people under SE declaration can easily migrate to nearby prefectures when they wished, to visit Japanese pubs and bars.

To avoid a potential underestimation of the impact, we excluded people who lived within 10 km and 20 km from the border in Columns (4) and (5), respectively. These results are consistent with our predictions. The estimated odds ratio (OR) for people who lived in prefectures with the SE declaration was 0.820 (CI95 0.700-0.960) in Column (4). The effect in terms of odds ratio is much higher in Column (5) (OR 0.771; CI95 0.640-0.930).

We move to the subsample analysis next because the early closure of Japanese pubs and bars has nothing to do with those who do not use them frequently. In fact, in SI Appendix Figure SI1, we find that the utilization rate of restaurants and bars was 48 % among people who were not scared of SARS-CoV-2 at all, but 30 % among those who were very scared in August-September 2020, when no restrictive measures were implemented.

With this in mind, we implemented a subsample analysis according to age, educational qualification, use of Japanese pubs and bars before the pandemic, and how scared of SARS-CoV-2. The results are presented in Figure 3. In this figure, the odds ratio and 95 percent confidence intervals from the three specifications used in Columns (3) to (5) in Table 1 are plotted. The full tables for the results in Figure 3 are shown in SI Appendix Tables S2-S9.

**Figure 3:**
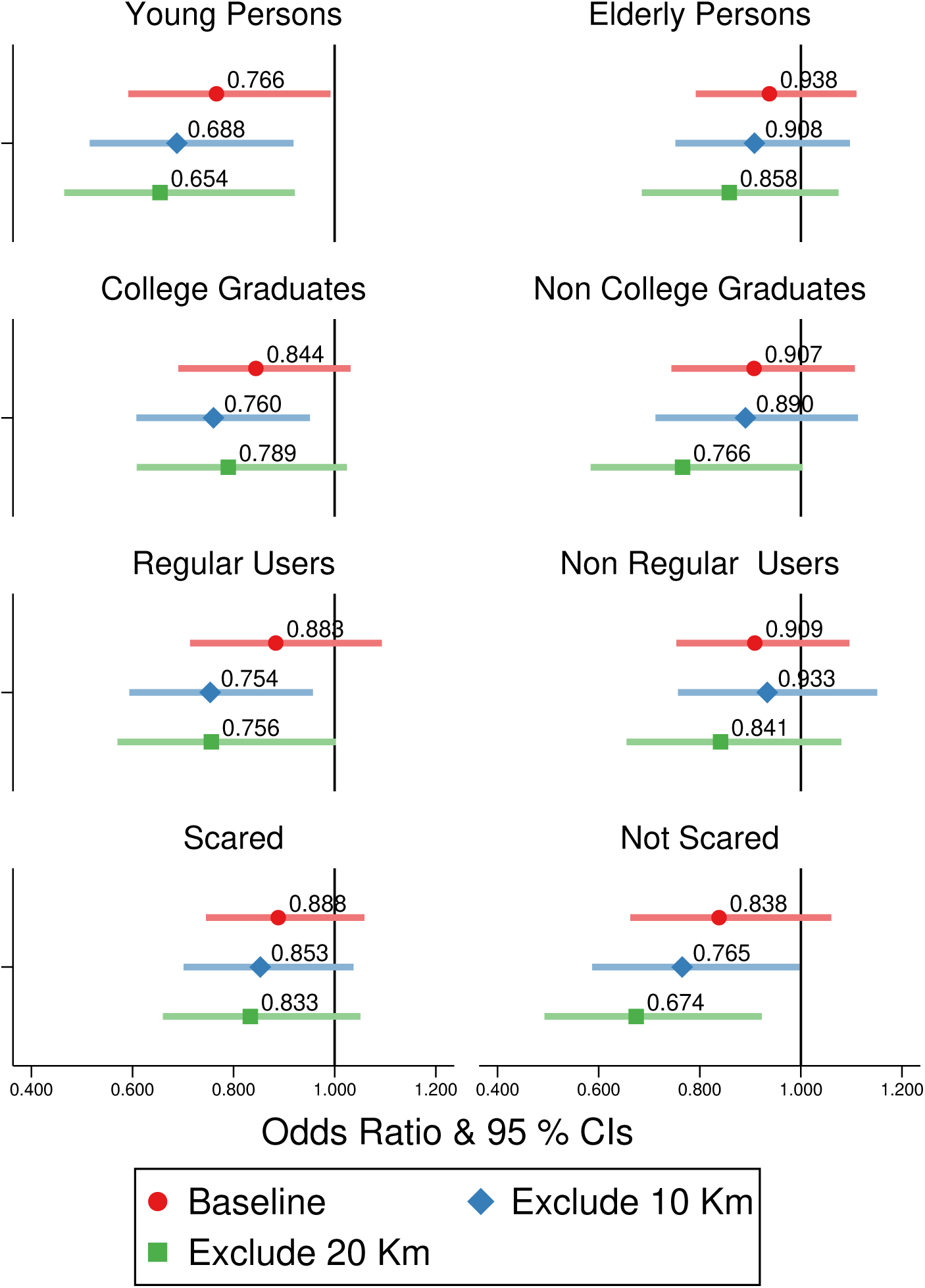
Effects on the utilization of Japanese pubs and bars by individual characteristics. *Notes:* Odds ratios (ORs) and 95 percent confidence intervals (CIs) were calculated using a random effect logistic regression model. The baseline specification controls for individual-level covariates as well as prefecture fixed effects, and excludes people who live in areas more than 50 km from the border of the prefectures with SE declarations. This specification is also used in Column (3) of Table 1. The “young persons” represents those aged less than 40 years old. “Regular users” represents the individuals who used Japanese pubs, which are a common type of full service restaurant in Japan, and bars before the pandemic. We control for age, working status (i.e., job loss or not), marital status, smoking, and household income level. The household income level is measured by a binary variable that takes a value of 1 for households with an annual income of JPY 7 million (USD 70,000) or more.

From Figure 3, we find that the utilization of Japanese pubs and bars reduced among young persons and regular users, while there are no statistically significant decreases among elderly persons and non-users. Among young people, the odds ratio was 0.688 (CI95: 0.515-0.918) in our preferred specification, which excludes an area within 10 km from the border. In terms of percentage point changes derived from the fixed-effect linear probability model in SI Appendix Table S2, it is equivalent to a 7.2% point reduction in the utilization rate (CI95: -12.2pp - -2.2pp), indicating a large reduction of the utilization rate.

Regarding educational attainment, we found a much larger reduction in the use among college graduates than among non-college graduates. Regarding fear of SARS-CoV-2, we found that the ORs were lower among people who were not scared of the virus than among those who were scared. In terms of percentage point changes, the utilization rate among persons who was not scared of SARS-CoV-2 decreased by 4.7% points (CI95: -9.3pp - - 0.1pp) in SI Appendix Table S8.

### 3.2 Symptoms indicative of SARS-CoV-2

Given that the early closure of Japanese pubs and bars significantly reduces their use among some high-risk subpopulations, such as young people, and the users of these services, this may lead to the reduction of symptoms related to SARS-CoV-2 mainly among these groups.

According to the descriptive statistics for five symptoms (fever, sore throat, cough, headache, and smell and taste disorder) in the SI Appendix Table S1, the proportion of people with each symptom was 1.8% (fever), 11.4% (sore throat), 13.1% (cough), headache(1.9%), and 1.2% (smell and taste disorder). We also found that smell and taste disorder, which has the highest specificity for the infection of SARS-CoV-2 among these five symptoms, increased from 0.8% in August-September 2020, to 1.3% in February 2021.

The results of the full sample for the five symptoms are listed in Table 2. The specifications in Columns (1) to (5) are the same as those in Table 1. In this table, we do not find any statistically significant reductions at the 95% significance level for all the symptoms. In our preferred specifications in Column (4), the odds ratio for fever exceeds 1, strongly rejecting the reduction (OR 1.222; CI95 0.697-2.141]. The odds ratios for headache, smell, and taste disorder were 0.966[CI95 0.493-1.894] and 0.808 (CI95 0.362-1.807), respectively.

**Table 2:** Effects on symptoms like SARS-CoV-2

|  | (1) | (2) | (3) | (4) | (5) |
| --- | --- | --- | --- | --- | --- |
| Panel A. Fixed Effect Linear Probability Model |  |  |  |  |  |
| High Fever | 0.003<br>(-0.001 - 0.007) | 0.003<br>(-0.001 - 0.008) | 0.002<br>(-0.004 - 0.008) | 0.003<br>(-0.004 - 0.011) | 0.005<br>(-0.004 - 0.014) |
| Sore Throat | 0.001<br>(-0.013 - 0.015) | 0.001<br>(-0.015 - 0.017) | -0.009<br>(-0.027 - 0.009) | -0.01<br>(-0.031 - 0.011) | -0.016<br>(-0.042 - 0.011) |
| Cough | -0.005<br>(-0.016 - 0.006) | -0.007<br>(-0.020 - 0.006) | -0.015*<br>(-0.033 - 0.003) | -0.017<br>(-0.040 - 0.005) | -0.018<br>(-0.044 - 0.008) |
| Headache | -0.020*<br>(-0.040 - 0.000) | -0.023*<br>(-0.047 - 0.001) | -0.02<br>(-0.054 - 0.013) | -0.011<br>(-0.048 - 0.025) | 0.005<br>(-0.045 - 0.054) |
| Smell and Taste Disorder | 0.002<br>(-0.003 - 0.006) | 0.001<br>(-0.003 - 0.005) | 0.001<br>(-0.002 - 0.005) | -0.001<br>(-0.004 - 0.003) | -0.001<br>(-0.005 - 0.003) |
| Panel B. Random Effect Logistic Regression Model |  |  |  |  |  |
| High Fever | 1.302<br>(0.919 - 1.843) | 1.258<br>(0.868 - 1.824) | 1.15<br>(0.697 - 1.896) | 1.222<br>(0.697 - 2.141) | 1.279<br>(0.670 - 2.444) |
| Sore Throat | 1.024<br>(0.881 - 1.191) | 1.03<br>(0.871 - 1.218) | 0.89<br>(0.713 - 1.110) | 0.894<br>(0.701 - 1.141) | 0.84<br>(0.631 - 1.118) |
| Cough | 0.97<br>(0.836 - 1.124) | 0.941<br>(0.798 - 1.109) | 0.844<br>(0.681 - 1.045) | 0.811*<br>(0.640 - 1.028) | 0.796<br>(0.598 - 1.058) |
| Headache | 0.797<br>(0.525 - 1.210) | 0.778<br>(0.495 - 1.221) | 0.937<br>(0.510 - 1.722) | 0.966<br>(0.493 - 1.894) | 0.987<br>(0.454 - 2.147) |
| Smell and Taste Disorder | 1.187<br>(0.741 - 1.903) | 1.095<br>(0.659 - 1.818) | 1.111<br>(0.546 - 2.262) | 0.808<br>(0.362 - 1.807) | 0.809<br>(0.331 - 1.980) |
| Prefecture fixed effects for logistic regression | yes | yes | yes | yes | yes |
| Individual-level covariates | No | Yes | Yes | Yes | Yes |
| Exclude Respondents far from 50 km from the Border | no | no | yes | yes | yes |
| Exclude Respondents within 10 Km from the Border | no | no | no | yes | no |
| Exclude Respondents within 20 Km from the Border | no | no | no | no | yes |
| Observations | 20,619 | 17,132 | 13,214 | 11,280 | 7,982 |

*Notes:* The dependent variable is a binary variable for the utilization of full service restaurants (Japanese pubs) and bars in the past one month. In Panel B, the results are based on a random effect logistic regression model. Odds ratios (ORs) and 95 confidence intervals (CIs) were reported. We control for age, working status (i.e., job loss or not), marital status, smoking, and household income level. The household income level is measured by a binary variable that takes a value of 1 for households with an annual income of JPY 7 million (USD 70,000) or more. Standard errors are reported in parentheses. \*\*\* p<0.01; \*\* p<0.05; \* p<0.1.

Figure 4 summarizes the results of the subsample analysis. Although we find large reductions in the utilization of Japanese pubs and bars among young people, regular users, and those who were not scared at SARS-CoV-2, the odds ratio for fever among young persons is 0.960[CI95 0.431 - 2.134], suggesting no statistically significant reduction. On smell and taste disorder, we find odds ratios are rather positive among young persons, regular users, and those who were not scared at SARS-CoV-2. Only among college graduates do we find a statistically significant reduction of cough (OR 0.651; CI95 0.465 - 0.911).

**Figure 4:**
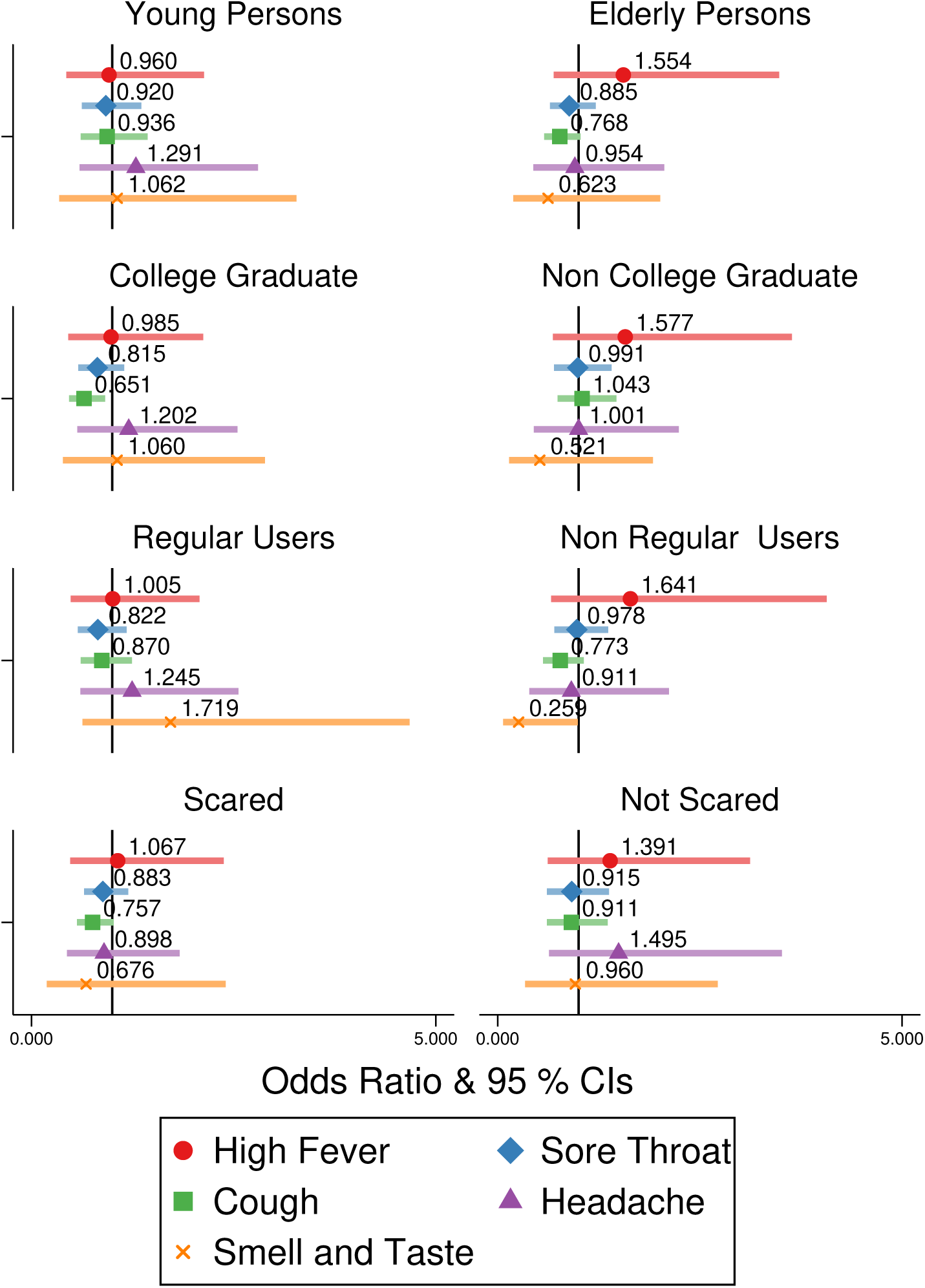
Effects on Symptoms of COVID-19 *Notes:* Odds ratios (ORs) and 95 percent confidence intervals (CIs) were calculated using the random effect logistic regression model. The specification is the same as that in Column (4) of Table 2. “Young persons” represents those less than 40 years old. “Regular users” represents individuals who regularly used Japanese pubs, which are a common type of full service restaurant in Japan, and bars before the pandemic. We control for age, working status (i.e., job loss or not), marital status, smoking, and household income level. The household income level is measured by a binary variable that takes a value of 1 for households with an annual income of JPY 7 million (USD 70,000) or more.

We found no statistically significant effects for the outcomes among other subpopulations. Among elderly persons who were not strongly affected by the strict measures for Japanese pubs and bars, all odds ratios were not statistically significant, as expected.

The results from the fixed-effect linear probability model in SI Appendix Tables S10-S17 is totally consistent with those from the random effect logistic regression model.

## 4 Discussion

In this paper, we explore how the early closure of full-service restaurants (Japanese pubs) and bars in Japan during the third wave of the SARS-CoV-2 pandemic affected the utilization of these services and the incidence of symptoms indicative of SARS-CoV-2. During the third wave, 11 prefectures declared an SE in January 2021 and prohibited restaurants and bars to serve alcoholic beverages after 7 p.m. and forced them to close by 8 p.m. Unlike other countries experiencing the first wave, no other strict measures such as school closures and border restrictions were implemented in Japan. Therefore, Japan during the winter 2020-2021 can be regarded as an ideal laboratory to explore the effects of the early closure of restaurants and bars.

By using large-scale nationally representative survey data and focusing on the people who live sufficiently close to the border that separates prefectures with and without SE declarations, we find a large reduction in the utilization of Japanese *Izakaya* pubs, which are a common type of full-service restaurants in Japan, and bars. In particular, the impacts are large among young people(OR 0.688; CI95 0.515 - 0.918) and regular users of these services (OR 0.754; CI95 0.594 - 0.957). Despite the reduction of the utilization rate, we find no discernible decrease in the symptoms of SARS-CoV-2, except the reduction of “cough” among college graduates. In the inactive and low-risk subpopulations (e.g., elderly persons), as expected, no changes were observed in both the utilization and the symptoms.

The results from our analysis exhibit a sharp contrast to the existing evidence that supports the effectiveness of the early closure of full-service restaurants and bars in restraining the spread of SARS-CoV-2 (Chang et al., 2020; Fisher et al., 2020; Brauner et al., 2021; Askitas, Tatsiramos and Verheyden, 2021; Haug et al., 2020; Persson, Parie and Feuerriegel, 2021). There are several reasons why our study did not find any discernible impacts of the early closure policies. First, it is possible that the effectiveness of the early closure varies according to the standard of daily preventive behaviors. In the second and later waves, most people knew the importance of wearing masks and washing hands frequently. In particular, compliance with these preventive behaviors was quite high in Japan because of its collectivist culture (Lu, Jin and English, 2021), which is different from that in the US (Bazzi, Fiszbein and Gebresilasse, 2021). Thus, it is possible that the additional effects of early closure, though not completely null, are limited. Therefore, more research is needed to investigate the effects of the early closure changes over time.

Second, early closure affects only high risk subpopulations, who may be prone to other risky behaviors. Thus, it is possible that they will eventually become infected, even if full-service restaurants and bars are closed. To support this interpretation, we additionally explore the effects on daily going-out behaviors such as “avoid crowds” in SI Appendix Figures S2. In short, we find no discernible impacts on these outcomes, suggesting that, even without going to full-service restaurants and bars, the overall tendency to go out remained almost unchanged among high-risk subpopulations. This is consistent with evidence from the UK that the early closure of full-service restaurants and bars in September 2020 did not reduce person-to-person contact (Jarvis et al., 2021).

Note that our results do not seem to be compatible with some evidence that government subsidies aimed at encouraging people to eat out in restaurants such as the UK’s Eat-Out-to-Help-Out scheme spread SARS-CoV-2 (Fetzer et al., 2020). One study in Japan also found that the Japanese version of this government subsidy increased the symptoms of SARS-CoV-2 (Miyawaki et al., 2021). However, it is natural to think that subsidization and early closure may have asymmetric impacts on the spread of SARS-CoV-2. Specifically, the subsidization to eat out in restaurants largely increased the number of people coming into contact with each other, but the early closure may have limited impacts on such contacts because people meet and chat with their friends and colleagues in other places (Jarvis et al., 2021). Therefore, our results are still compatible with earlier studies on the effects of subsidies (Fetzer et al., 2020; Miyawaki et al., 2021).

Although this study is the first to investigate the independent causal effects of the early closure of restaurants and bars, there are several limitations. First, in our experimental setting, individuals in the prefectures with SE declarations could migrate to nearby prefectures to eat in restaurants and drink at bars. Thus, it is still possible that a nationwide closure, not local closures, would suppress the spread of SARS-CoV-2. However, in the preferred specification used in Figure 4, individuals living in an area within 10 km from the border to separate prefectures with and without SE declarations were excluded because they can easily cross the border; thus, our results are sufficiently robust to potential migration, and implications from our paper can be still applicable for the effects of a nationwide closure.

The second limitation is related to the stable unit treatment value assumption (SUTVA) (Rubin, 2005; Halloran & Struchiner, 1995; VanderWeele, 2008). Because infectious diseases spread locally and rapidly, neighboring areas generally tend to have similar infection rates. Therefore, our research may underestimate the impact of the policy, even if the early closure of restaurants and bars suppresses SARS-CoV-2 to a large extend. However, if this threat is relevant, we should have found large negative effects in the models that include persons far from the border, like in Columns (1) and (2) in Table 2, because the spillover effects from the prefectures with SE declarations to those without may not reach areas far away from the border. However, the odds ratios from these specifications regarding fever, sore throat, cough, and smell and taste disorder are generally higher than the odds ratios in Columns (3) to (4). This pattern of results is not compatible with the instantaneous spread of SARS-CoV-2 within local areas. Previous studies have also shown that SARS-CoV-2 spreads through commuting (Kissler et al., 2020) and traveling (Chinazzi et al., 2020), which are not always limited to areas that are geographically close to each other.

From the perspective of public policy implications, our study suggests that the early closure of full-service restaurants and bars, without any other concurrent policies, is not an efficient way to suppress SARS-CoV-2. Given the large detrimental effects on employment (Kong & Prinz, 2020; Miyakawa, Oikawa and Ueda, 2021), alternative measures for full-service restaurants and bars should be considered before they are closed completely. For example, a combination of much more moderate NPIs, such as bans on small gatherings (Persson, Parie and Feuerriegel, 2021) and early detection and isolation (Lai et al., 2020) would perform better than only targeting full-service restaurants and bars. Evidence also suggests that the effectiveness of NPIs largely depends on the combination of the interventions implemented (Nishi et al., 2020; Haug et al., 2020; Brauner et al., 2021). In addition, in an effective combination with other NPIs and a sufficiently high vaccination rate today, the early closure policy can be replaced by weaker measures, such as a limitation on the number of seats served per table. Such moderate interventions would be, if implemented successfully, far more efficient to make the infection control compatible with the freedom of business among full-service restaurants and bars.

## Data Availability

Data are prohibited to be shared due to the confidential nature.

## Supplementary Information Appendix

**Table SI1:** Descriptive Statistics

|  | Full Sample | Prefectures<br>with the SE | Prefectures<br>without the SE |
| --- | --- | --- | --- |
|  | (1)<br>Mean<br>(S.D.) | (2)<br>Mean<br>(S.D.) | (3)<br>Mean<br>(S.D.) |
| **Dependent Variables** |  |  |  |
| Utilization of Restaurants and Bars During Past One Month | 0.378<br>(0.485) | 0.387<br>(0.487) | 0.351<br>(0.477) |
| Fever | 0.0184<br>(0.135) | 0.0192<br>(0.137) | 0.0161<br>(0.126) |
| Sore Throat | 0.114<br>(0.317) | 0.114<br>(0.318) | 0.112<br>(0.316) |
| Cough | 0.131<br>(0.337) | 0.129<br>(0.335) | 0.136<br>(0.343) |
| Headache | 0.0187<br>(0.135) | 0.0198<br>(0.139) | 0.0155<br>(0.123) |
| Smell and Taste Disorder | 0.0122<br>(0.110) | 0.0125<br>(0.111) | 0.0114<br>(0.106) |
| **Individual Characteristics** |  |  |  |
| Age | 50.53<br>(16.510) | 49.92<br>(16.600) | 52.34<br>(16.100) |
| College Graduate | 0.512<br>(0.500) | 0.531<br>(0.499) | 0.455<br>(0.498) |
| Married | 0.629<br>(0.483) | 0.615<br>(0.487) | 0.67<br>(0.470) |
| Worker | 0.642<br>(0.480) | 0.648<br>(0.478) | 0.622<br>(0.485) |
| Household with Income More Than JPY 7 Million | 0.325<br>(0.468) | 0.338<br>(0.473) | 0.285<br>(0.452) |
| Smoker | 0.21<br>(0.407) | 0.212<br>(0.408) | 0.205<br>(0.403) |
| Used Japanese Pubs and Bars During the Period Before the Pandemic<br>("Japanese Pub Users") | 0.456<br>(0.498) | 0.474<br>(0.499) | 0.401<br>(0.490) |
| Not Scared at SARS-CoV-2 | 0.373<br>(0.484) | 0.379<br>(0.485) | 0.356<br>(0.479) |
| Observations | 25,338 | 19,007 | 6,331 |

*Notes:* The sample consists of persons who lived within 50 Km from the border which separates the prefectures with and without the SE (state of emergency) declaration.

**Figure SI1:**
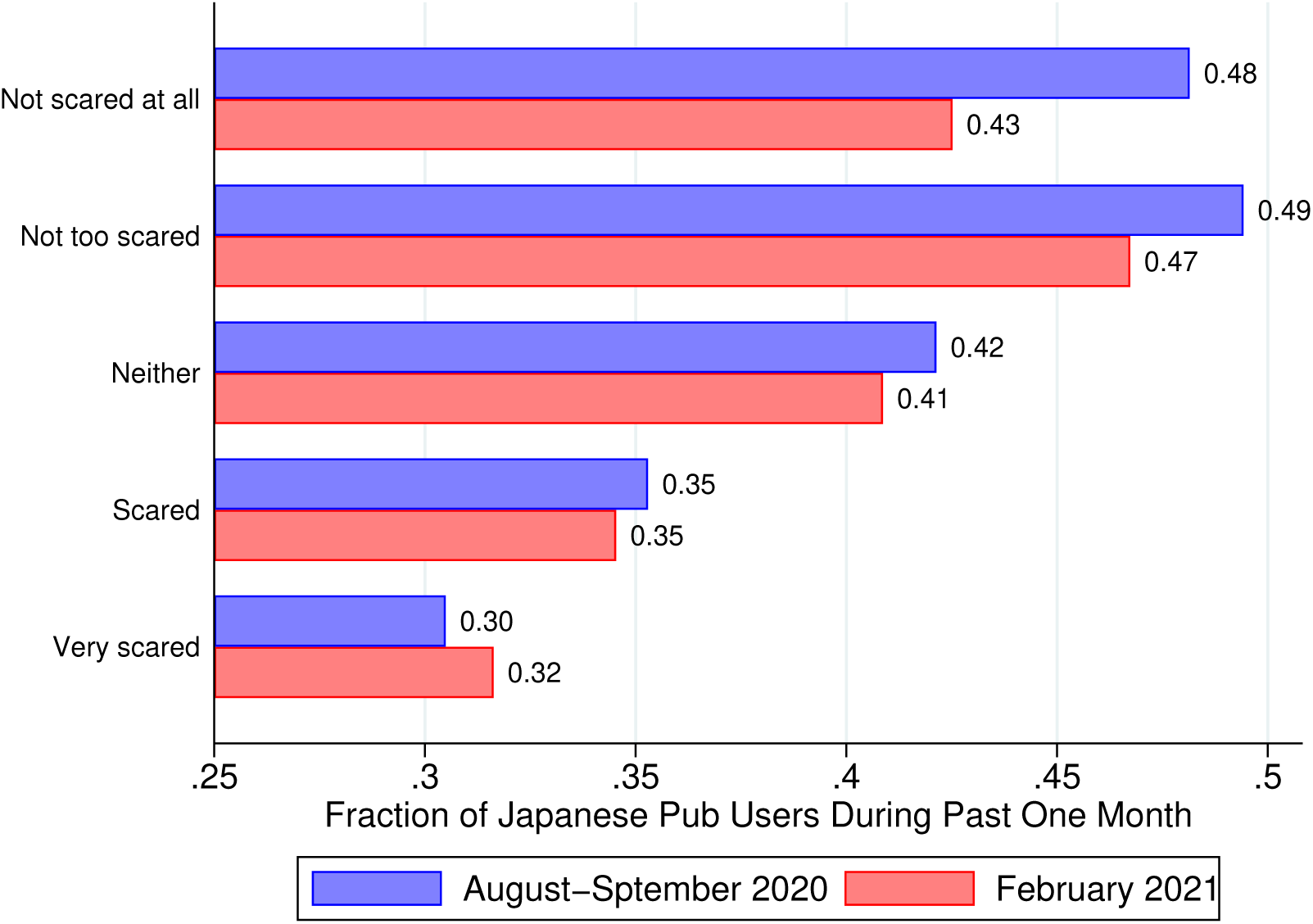
Utilization Rate of Japanese Pubs and Bars by the Extent of Fear of SARS-CoV-2

**Table SI2:**
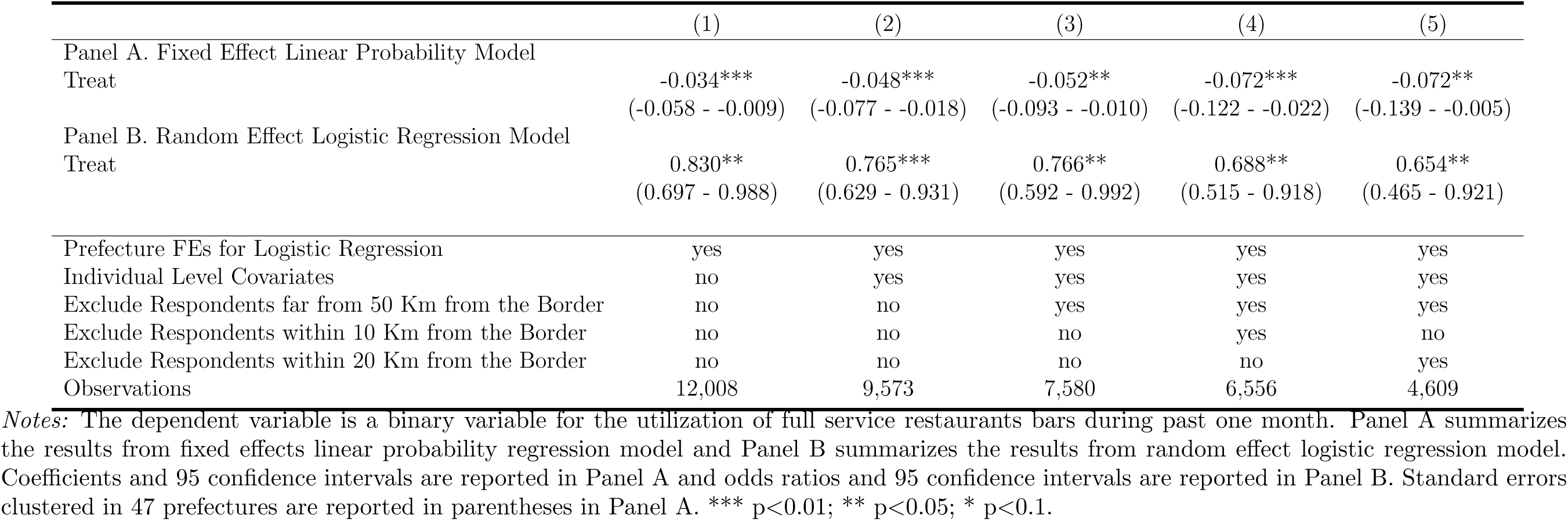
Effects on the Utilization of Japanese Pubs and Bars among Young Persons

**Table SI3:**
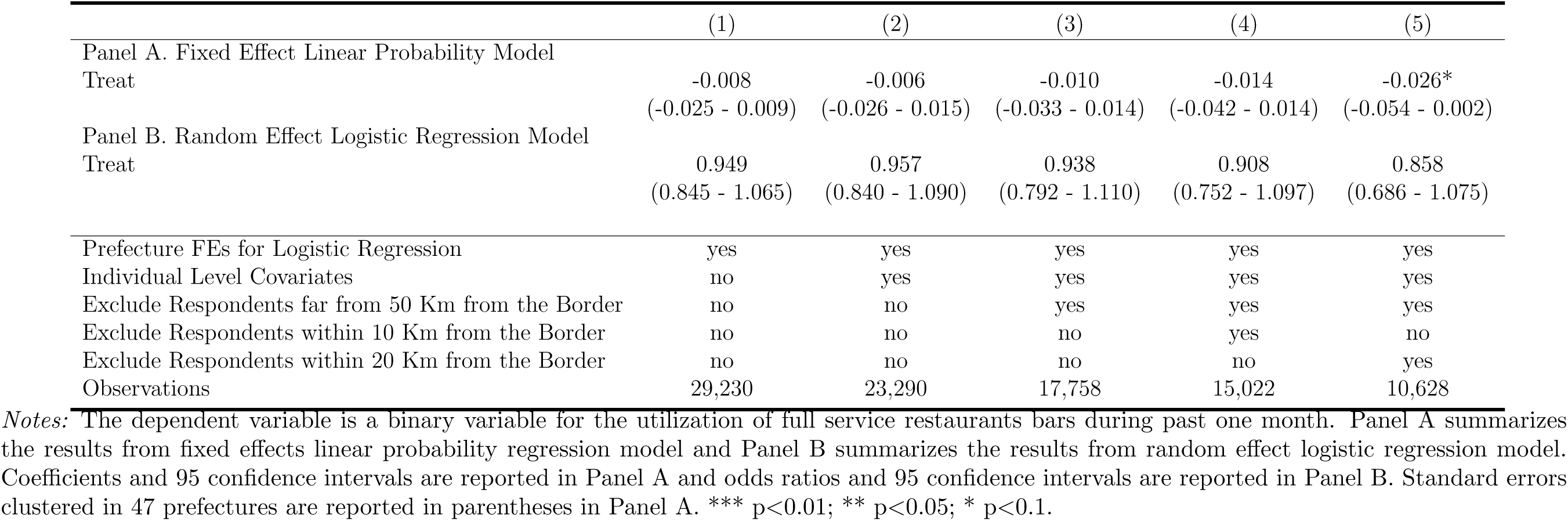
Effects on the Utilization of Japanese Pubs and Bars among Elderly Persons

**Table SI4:** Effects on the Utilization of Japanese Pubs and Bars among College Graduates

|  | (1) | (2) | (3) | (4) | (5) |
| --- | --- | --- | --- | --- | --- |
| Panel A. Fixed Effect Linear Probability Model |  |  |  |  |  |
| Treat | -0.031***<br>(-0.053 - -0.009) | -0.037***<br>(-0.063 - -0.011) | -0.033*<br>(-0.068 - 0.002) | -0.048**<br>(-0.089 - -0.007) | -0.046*<br>(-0.093 - 0.001) |
| Panel B. Random Effect Logistic Regression Model |  |  |  |  |  |
| Treat | 0.832**<br>(0.722 - 0.957) | 0.800***<br>(0.685 - 0.935) | 0.844*<br>(0.691 - 1.031) | 0.760**<br>(0.608 - 0.951) | 0.789*<br>(0.608 - 1.024) |
| Prefecture FEs for Logistic Regression | yes | yes | yes | yes | yes |
| Individual Level Covariates | no | yes | yes | yes | yes |
| Exclude Respondents far from 50 Km from the Border | no | no | yes | yes | yes |
| Exclude Respondents within 10 Km from the Border | no | no | no | yes | no |
| Exclude Respondents within 20 Km from the Border | no | no | no | no | yes |
| Observations | 20,017 | 16,581 | 13,427 | 11,630 | 8,452 |

*Notes:* The dependent variable is a binary variable for the utilization of full service restaurants bars during past one month. Panel A summarizes the results from fixed effects linear probability regression model and Panel B summarizes the results from random effect logistic regression model. Coefficients and 95 confidence intervals are reported in Panel A and odds ratios and 95 confidence intervals are reported in Panel B. Standard errors clustered in 47 prefectures are reported in parentheses in Panel A. \*\*\* p<0.01; \*\* p<0.05; \* p<0.1.

**Table SI5:** Effects on the Utilization of Japanese Pubs and Bars among Non College Graduates

|  | (1) | (2) | (3) | (4) | (5) |
| --- | --- | --- | --- | --- | --- |
| Panel A. Fixed Effect Linear Probability Model |  |  |  |  |  |
| Treat | -0.009<br>(-0.030 - 0.013) | -0.005<br>(-0.029 - 0.019) | -0.013<br>(-0.044 - 0.017) | -0.019<br>(-0.055 - 0.018) | -0.039*<br>(-0.079 - 0.002) |
| Panel B. Random Effect Logistic Regression Model |  |  |  |  |  |
| Treat | 0.951<br>(0.832 - 1.088) | 0.970<br>(0.832 - 1.131) | 0.907<br>(0.744 - 1.107) | 0.890<br>(0.712 - 1.113) | 0.766*<br>(0.584 - 1.004) |
| Prefecture FEs for Logistic Regression | yes | yes | yes | yes | yes |
| Individual Level Covariates | no | yes | yes | yes | yes |
| Exclude Respondents far from 50 Km from the Border | no | no | yes | yes | yes |
| Exclude Respondents within 10 Km from the Border | no | no | no | yes | no |
| Exclude Respondents within 20 Km from the Border | no | no | no | no | yes |
| Observations | 21,131 | 16,229 | 11,876 | 9,914 | 6,755 |

*Notes:* The dependent variable is a binary variable for the utilization of full service restaurants bars during past one month. Panel A summarizes the results from fixed effects linear probability regression model and Panel B summarizes the results from random effect logistic regression model. Coefficients and 95 confidence intervals are reported in Panel A and odds ratios and 95 confidence intervals are reported in Panel B. Standard errors clustered in 47 prefectures are reported in parentheses in Panel A. \*\*\* p<0.01; \*\* p<0.05; \* p<0.1.

**Table SI6:** Effects on the Utilization of Japanese Pubs and Bars among Japanese Pub Users

|  | (1) | (2) | (3) | (4) | (5) |
| --- | --- | --- | --- | --- | --- |
| Panel A. Fixed Effect Linear Probability Model |  |  |  |  |  |
| Treat | -0.016<br>(-0.043 - 0.012) | -0.023<br>(-0.052 - 0.006) | -0.027<br>(-0.069 - 0.015) | -0.054**<br>(-0.096 - -0.013) | -0.053*<br>(-0.109 - 0.003) |
| Panel B. Random Effect Logistic Regression Model |  |  |  |  |  |
| Treat | 0.914<br>(0.788 - 1.060) | 0.882<br>(0.751 - 1.037) | 0.883<br>(0.714 - 1.093) | 0.754**<br>(0.594 - 0.957) | 0.756*<br>(0.570 - 1.002) |
| Prefecture FEs for Logistic Regression | yes | yes | yes | yes | yes |
| Individual Level Covariates | no | yes | yes | yes | yes |
| Exclude Respondents far from 50 Km from the Border | no | no | yes | yes | yes |
| Exclude Respondents within 10 Km from the Border | no | no | no | yes | no |
| Exclude Respondents within 20 Km from the Border | no | no | no | no | yes |
| Observations | 17,282 | 14,697 | 11,550 | 9,972 | 7,124 |

*Notes:* The dependent variable is a binary variable for the utilization of full service restaurants bars during past one month. Panel A summarizes the results from fixed effects linear probability regression model and Panel B summarizes the results from random effect logistic regression model. Coefficients and 95 confidence intervals are reported in Panel A and odds ratios and 95 confidence intervals are reported in Panel B. Standard errors clustered in 47 prefectures are reported in parentheses in Panel A. \*\*\* p<0.01; \*\* p<0.05; \* p<0.1.

**Table SI7:**
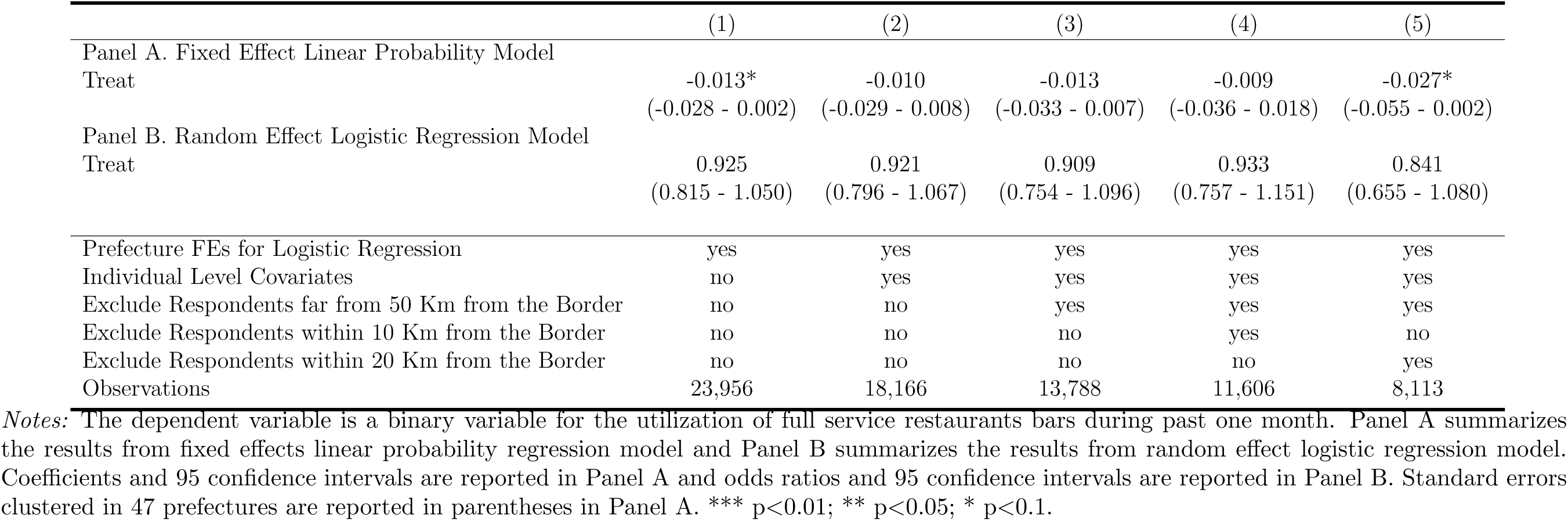
Effects on the Utilization of Japanese Pubs and Bars among Non Japanese Pub Users

**Table SI8:**
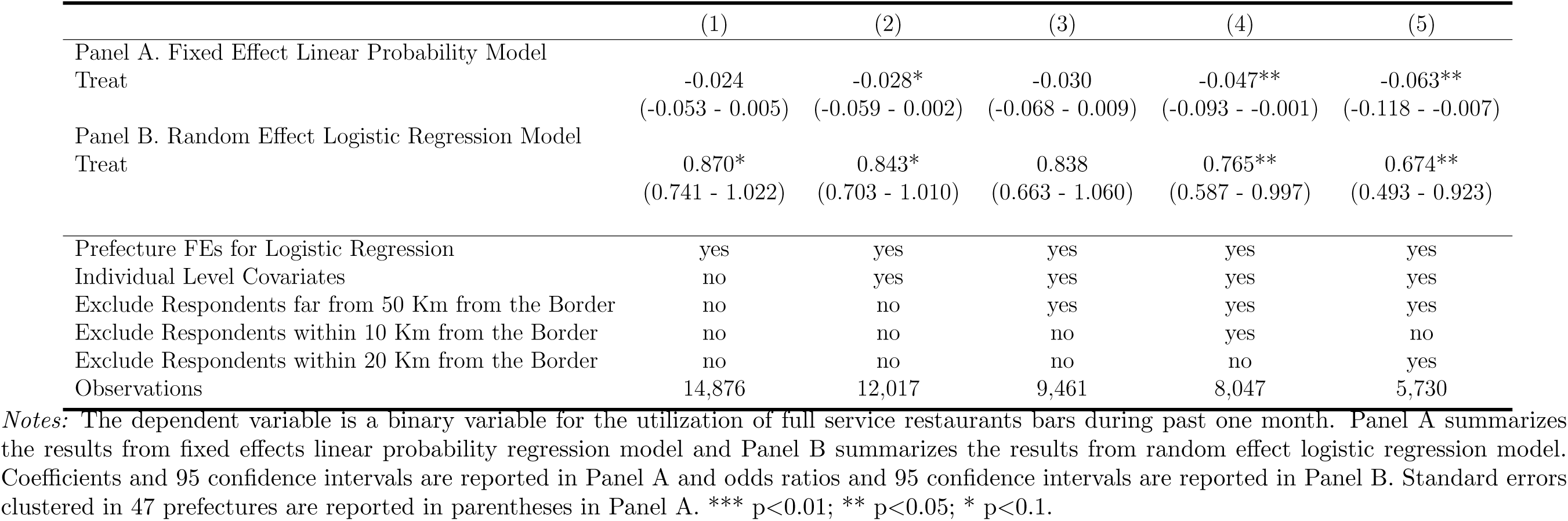
Effects on the Utilization of Japanese Pubs and Bars among ”Not Scared”

**Table SI9:**
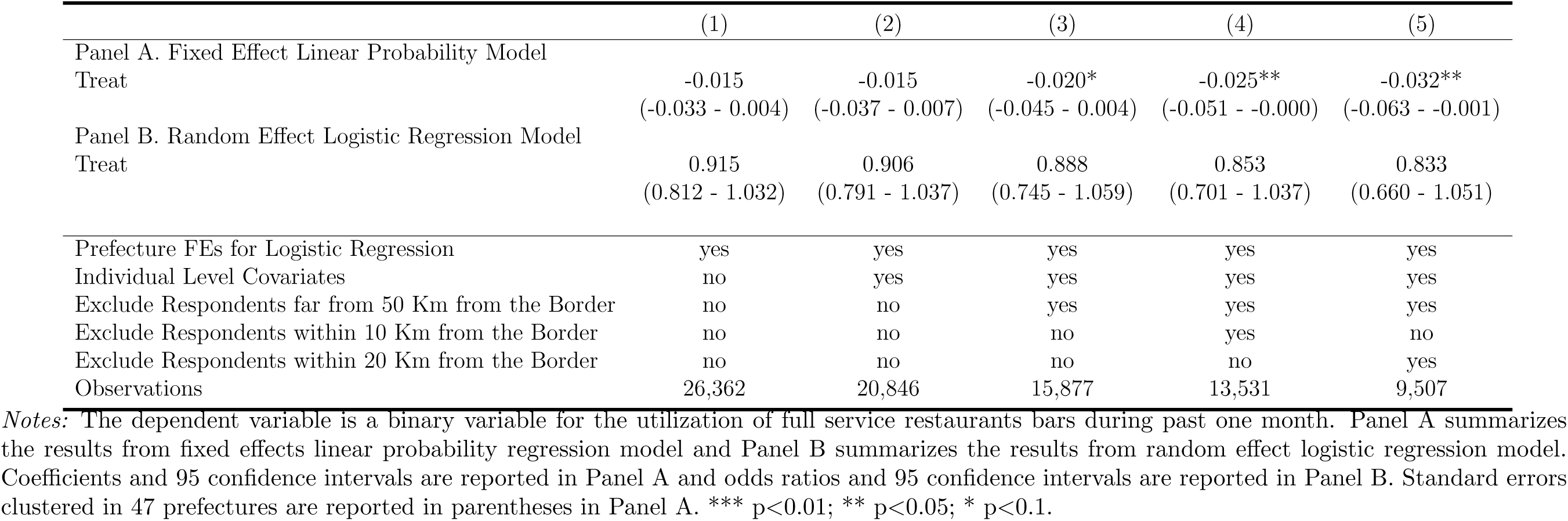
Effects on the Utilization of Japanese Pubs and Bars among “Scared”

**Table SI10:**
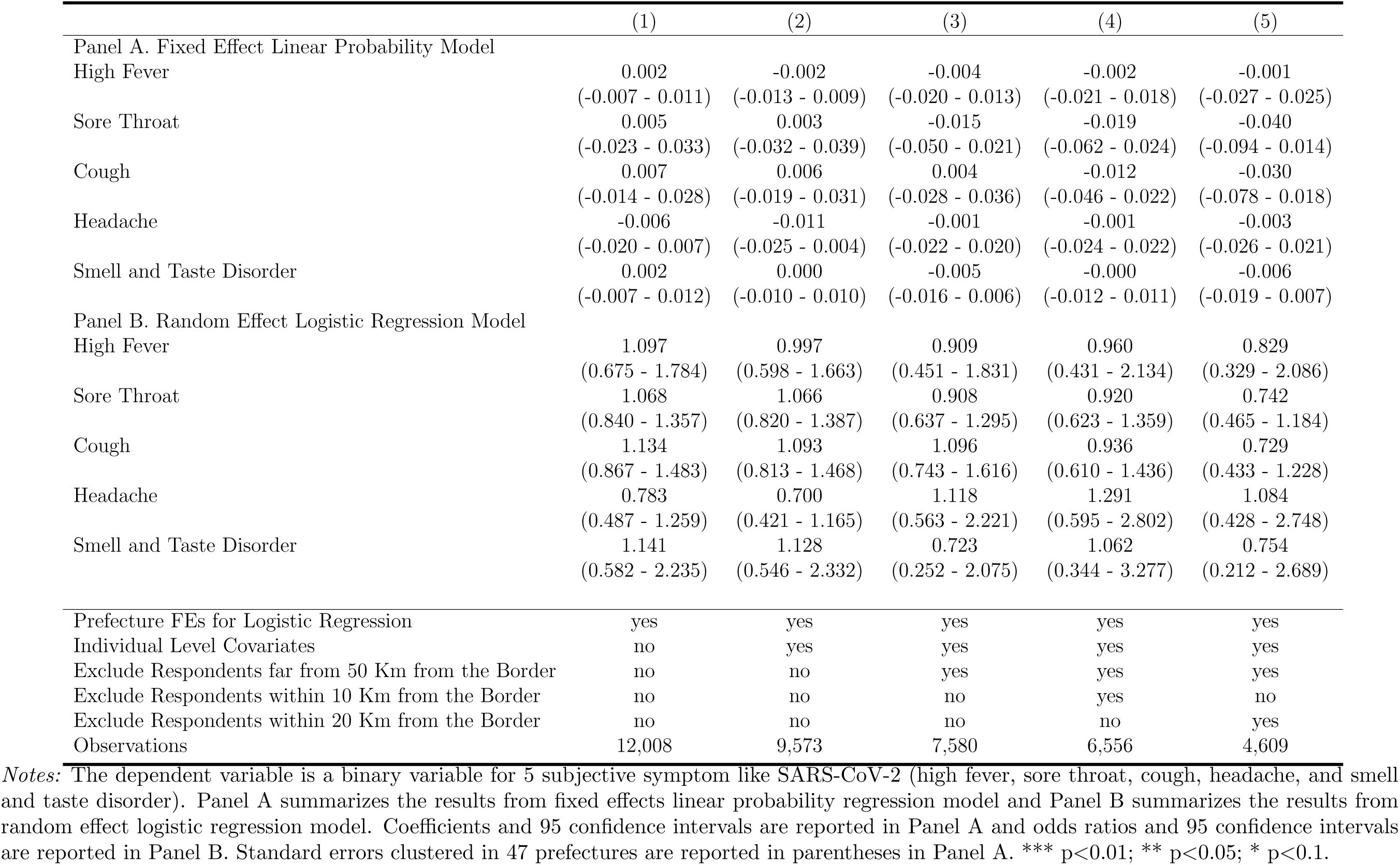
Effects on Symptoms Like SARS-CoV-2 among Young Persons

**Table SI11:**
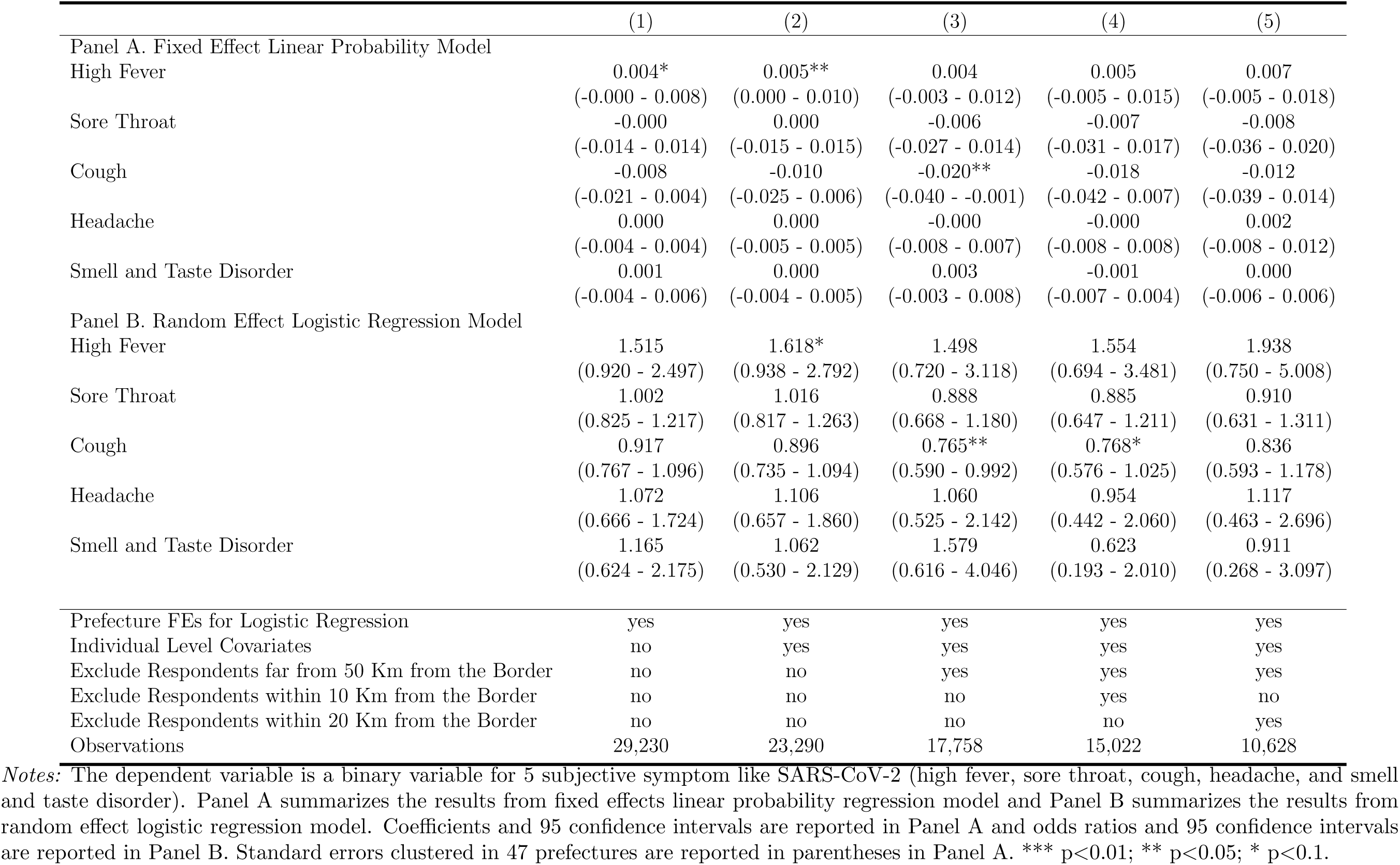
Effects on Symptoms Like SARS-CoV-2 among Elderly Persons

**Table SI12:** Effects on Symptoms Like SARS-CoV-2 among College Graduates

|  | (1) | (2) | (3) | (4) | (5) |
| --- | --- | --- | --- | --- | --- |
| Panel A. Fixed Effect Linear Probability Model |  |  |  |  |  |
| High Fever | 0.000<br>(-0.006 - 0.007) | -0.001<br>(-0.009 - 0.006) | 0.001<br>(-0.006 - 0.007) | 0.002<br>(-0.005 - 0.009) | 0.003<br>(-0.006 - 0.011) |
| Sore Throat | -0.010<br>(-0.029 - 0.009) | -0.009<br>(-0.029 - 0.011) | -0.014<br>(-0.037 - 0.009) | -0.017<br>(-0.045 - 0.011) | -0.014<br>(-0.051 - 0.023) |
| Cough | -0.013<br>(-0.031 - 0.005) | -0.016*<br>(-0.036 - 0.003) | -0.032***<br>(-0.055 - -0.008) | -0.036**<br>(-0.067 - -0.006) | -0.030<br>(-0.066 - 0.006) |
| Headache | -0.002<br>(-0.010 - 0.006) | -0.003<br>(-0.011 - 0.006) | -0.000<br>(-0.013 - 0.013) | 0.003<br>(-0.012 - 0.018) | 0.005<br>(-0.013 - 0.023) |
| Smell and Taste Disorder | 0.002<br>(-0.004 - 0.008) | 0.001<br>(-0.005 - 0.008) | 0.002<br>(-0.003 - 0.008) | 0.001<br>(-0.005 - 0.008) | 0.002<br>(-0.003 - 0.008) |
| Panel B. Random Effect Logistic Regression Model |  |  |  |  |  |
| High Fever | 0.951<br>(0.587 - 1.541) | 0.869<br>(0.523 - 1.444) | 1.001<br>(0.508 - 1.971) | 0.985<br>(0.456 - 2.124) | 0.939<br>(0.404 - 2.183) |
| Sore Throat | 0.865<br>(0.692 - 1.082) | 0.894<br>(0.702 - 1.138) | 0.838<br>(0.614 - 1.143) | 0.815<br>(0.579 - 1.147) | 0.827<br>(0.555 - 1.231) |
| Cough | 0.842<br>(0.678 - 1.048) | 0.815*<br>(0.644 - 1.031) | 0.687**<br>(0.507 - 0.930) | 0.651**<br>(0.465 - 0.911) | 0.669**<br>(0.451 - 0.991) |
| Headache | 0.803<br>(0.498 - 1.293) | 0.784<br>(0.471 - 1.304) | 1.036<br>(0.515 - 2.081) | 1.202<br>(0.567 - 2.548) | 1.040<br>(0.448 - 2.414) |
| Smell and Taste Disorder | 1.227<br>(0.640 - 2.354) | 1.109<br>(0.562 - 2.187) | 1.143<br>(0.456 - 2.864) | 1.060<br>(0.389 - 2.888) | 1.215<br>(0.391 - 3.778) |
| Prefecture FEs for Logistic Regression | yes | yes | yes | yes | yes |
| Individual Level Covariates | no | yes | yes | yes | yes |
| Exclude Respondents far from 50 Km from the Border | no | no | yes | yes | yes |
| Exclude Respondents within 10 Km from the Border | no | no | no | yes | no |
| Exclude Respondents within 20 Km from the Border | no | no | no | no | yes |
| Observations | 20,017 | 16,581 | 13,427 | 11,630 | 8,452 |

*Notes:* The dependent variable is a binary variable for 5 subjective symptom like SARS-CoV-2 (high fever, sore throat, cough, headache, and smell and taste disorder). Panel A summarizes the results from fixed effects linear probability regression model and Panel B summarizes the results from random effect logistic regression model. Coefficients and 95 confidence intervals are reported in Panel A and odds ratios and 95 confidence intervals are reported in Panel B. Standard errors clustered in 47 prefectures are reported in parentheses in Panel A. \*\*\* p<0.01; \*\* p<0.05; \* p<0.1.

**Table SI13:** Effects on Symptoms Like SARS-CoV-2 among Non College Graduates

|  | (1) | (2) | (3) | (4) | (5) |
| --- | --- | --- | --- | --- | --- |
| Panel A. Fixed Effect Linear Probability Model |  |  |  |  |  |
| High Fever | 0.005*<br>(-0.000 - 0.011) | 0.007*<br>(-0.001 - 0.014) | 0.002<br>(-0.009 - 0.014) | 0.002<br>(-0.012 - 0.016) | 0.003<br>(-0.011 - 0.018) |
| Sore Throat | 0.011<br>(-0.004 - 0.025) | 0.010<br>(-0.007 - 0.027) | -0.004<br>(-0.028 - 0.019) | -0.003<br>(-0.030 - 0.024) | -0.019<br>(-0.048 - 0.011) |
| Cough | 0.005<br>(-0.009 - 0.018) | 0.007<br>(-0.010 - 0.023) | 0.003<br>(-0.015 - 0.022) | 0.005<br>(-0.018 - 0.027) | -0.003<br>(-0.033 - 0.027) |
| Headache | 0.001<br>(-0.004 - 0.006) | -0.001<br>(-0.008 - 0.006) | 0.001<br>(-0.009 - 0.011) | -0.002<br>(-0.013 - 0.009) | -0.001<br>(-0.013 - 0.012) |
| Smell and Taste Disorder | 0.001<br>(-0.004 - 0.006) | 0.000<br>(-0.005 - 0.005) | 0.001<br>(-0.004 - 0.006) | -0.003<br>(-0.009 - 0.003) | -0.005<br>(-0.012 - 0.003) |
| Panel B. Random Effect Logistic Regression Model |  |  |  |  |  |
| High Fever | 1.778**<br>(1.070 - 2.955) | 1.919**<br>(1.102 - 3.343) | 1.412<br>(0.668 - 2.983) | 1.577<br>(0.684 - 3.637) | 1.732<br>(0.620 - 4.842) |
| Sore Throat | 1.190<br>(0.966 - 1.467) | 1.194<br>(0.941 - 1.515) | 0.957<br>(0.695 - 1.319) | 0.991<br>(0.697 - 1.409) | 0.834<br>(0.546 - 1.272) |
| Cough | 1.115<br>(0.907 - 1.370) | 1.126<br>(0.890 - 1.424) | 1.064<br>(0.782 - 1.448) | 1.043<br>(0.742 - 1.467) | 0.966<br>(0.634 - 1.471) |
| Headache | 1.074<br>(0.661 - 1.744) | 1.020<br>(0.598 - 1.742) | 1.185<br>(0.584 - 2.406) | 1.001<br>(0.447 - 2.240) | 1.110<br>(0.407 - 3.023) |
| Smell and Taste Disorder | 1.187<br>(0.614 - 2.296) | 1.072<br>(0.493 - 2.330) | 1.060<br>(0.358 - 3.142) | 0.521<br>(0.141 - 1.919) | 0.386<br>(0.084 - 1.783) |
| Prefecture FEs for Logistic Regression | yes | yes | yes | yes | yes |
| Individual Level Covariates | no | yes | yes | yes | yes |
| Exclude Respondents far from 50 Km from the Border | no | no | yes | yes | yes |
| Exclude Respondents within 10 Km from the Border | no | no | no | yes | no |
| Exclude Respondents within 20 Km from the Border | no | no | no | no | yes |
| Observations | 21,131 | 16,229 | 11,876 | 9,914 | 6,755 |

*Notes:* The dependent variable is a binary variable for 5 subjective symptom like SARS-CoV-2 (high fever, sore throat, cough, headache, and smell and taste disorder). Panel A summarizes the results from fixed effects linear probability regression model and Panel B summarizes the results from random effect logistic regression model. Coefficients and 95 confidence intervals are reported in Panel A and odds ratios and 95 confidence intervals are reported in Panel B. Standard errors clustered in 47 prefectures are reported in parentheses in Panel A. \*\*\* p<0.01; \*\* p<0.05; \* p<0.1.

**Table SI14:**
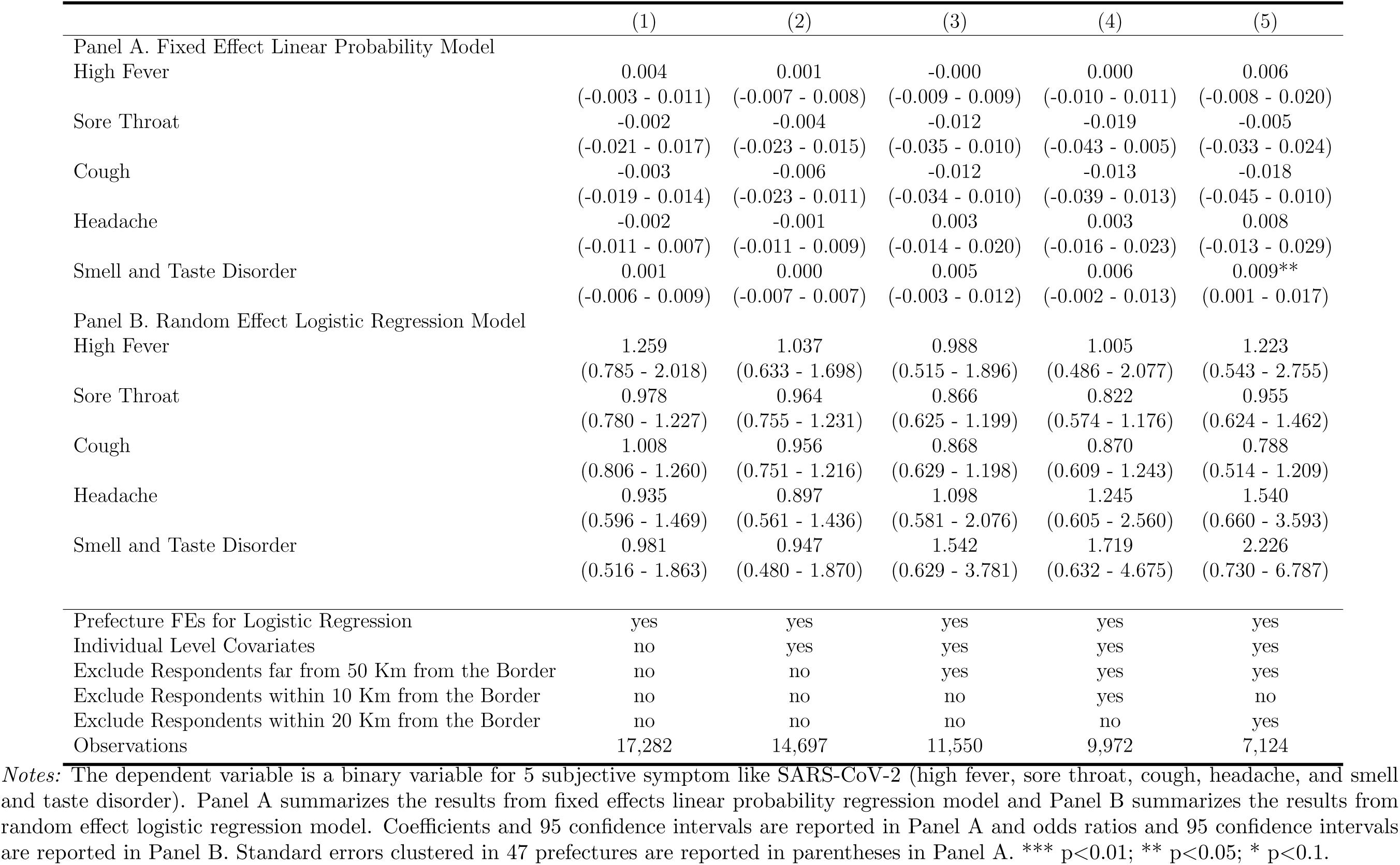
Effects on Symptoms Like SARS-CoV-2 among Regular Users

**Table SI15:** Effects on Symptoms Like SARS-CoV-2 among Non Regular Users

|  | (1) | (2) | (3) | (4) | (5) |
| --- | --- | --- | --- | --- | --- |
| Panel A. Fixed Effect Linear Probability Model |  |  |  |  |  |
| High Fever | 0.002<br>(-0.003 - 0.008) | 0.005<br>(-0.002 - 0.011) | 0.003<br>(-0.005 - 0.012) | 0.005<br>(-0.006 - 0.015) | 0.003<br>(-0.008 - 0.015) |
| Sore Throat | 0.005<br>(-0.011 - 0.020) | 0.006<br>(-0.012 - 0.024) | -0.005<br>(-0.027 - 0.017) | -0.003<br>(-0.028 - 0.022) | -0.023<br>(-0.057 - 0.010) |
| Cough | -0.006<br>(-0.018 - 0.006) | -0.006<br>(-0.024 - 0.011) | -0.017<br>(-0.041 - 0.007) | -0.020<br>(-0.052 - 0.013) | -0.017<br>(-0.050 - 0.016) |
| Headache | -0.001<br>(-0.006 - 0.004) | -0.004<br>(-0.010 - 0.002) | -0.003<br>(-0.013 - 0.006) | -0.004<br>(-0.015 - 0.007) | -0.005<br>(-0.018 - 0.008) |
| Smell and Taste Disorder | 0.001<br>(-0.003 - 0.005) | 0.001<br>(-0.004 - 0.005) | -0.002<br>(-0.008 - 0.005) | -0.006*<br>(-0.012 - 0.000) | -0.010***<br>(-0.016 - -0.003) |
| Panel B. Random Effect Logistic Regression Model |  |  |  |  |  |
| High Fever | 1.286<br>(0.765 - 2.161) | 1.583<br>(0.891 - 2.812) | 1.420<br>(0.644 - 3.134) | 1.641<br>(0.662 - 4.067) | 1.395<br>(0.467 - 4.170) |
| Sore Throat | 1.080<br>(0.882 - 1.323) | 1.110<br>(0.881 - 1.399) | 0.929<br>(0.687 - 1.258) | 0.978<br>(0.701 - 1.365) | 0.765<br>(0.517 - 1.130) |
| Cough | 0.945<br>(0.776 - 1.152) | 0.936<br>(0.747 - 1.173) | 0.830<br>(0.622 - 1.107) | 0.773<br>(0.561 - 1.065) | 0.799<br>(0.545 - 1.172) |
| Headache | 0.901<br>(0.544 - 1.492) | 0.839<br>(0.473 - 1.487) | 1.028<br>(0.473 - 2.236) | 0.911<br>(0.392 - 2.118) | 0.677<br>(0.246 - 1.866) |
| Smell and Taste Disorder | 1.245<br>(0.639 - 2.423) | 1.168<br>(0.543 - 2.510) | 0.673<br>(0.227 - 1.992) | 0.259**<br>(0.068 - 0.991) | 0.125**<br>(0.024 - 0.657) |
| Prefecture FEs for Logistic Regression | yes | yes | yes | yes | yes |
| Individual Level Covariates | no | yes | yes | yes | yes |
| Exclude Respondents far from 50 Km from the Border | no | no | yes | yes | yes |
| Exclude Respondents within 10 Km from the Border | no | no | no | yes | no |
| Exclude Respondents within 20 Km from the Border | no | no | no | no | yes |
| Observations | 23,956 | 18,166 | 13,788 | 11,606 | 8,113 |

*Notes:* The dependent variable is a binary variable for 5 subjective symptom like SARS-CoV-2 (high fever, sore throat, cough, headache, and smell and taste disorder). Panel A summarizes the results from fixed effects linear probability regression model and Panel B summarizes the results from random effect logistic regression model. Coefficients and 95 confidence intervals are reported in Panel A and odds ratios and 95 confidence intervals are reported in Panel B. Standard errors clustered in 47 prefectures are reported in parentheses in Panel A. \*\*\* p<0.01; \*\* p<0.05; \* p<0.1.

**Table SI16:**
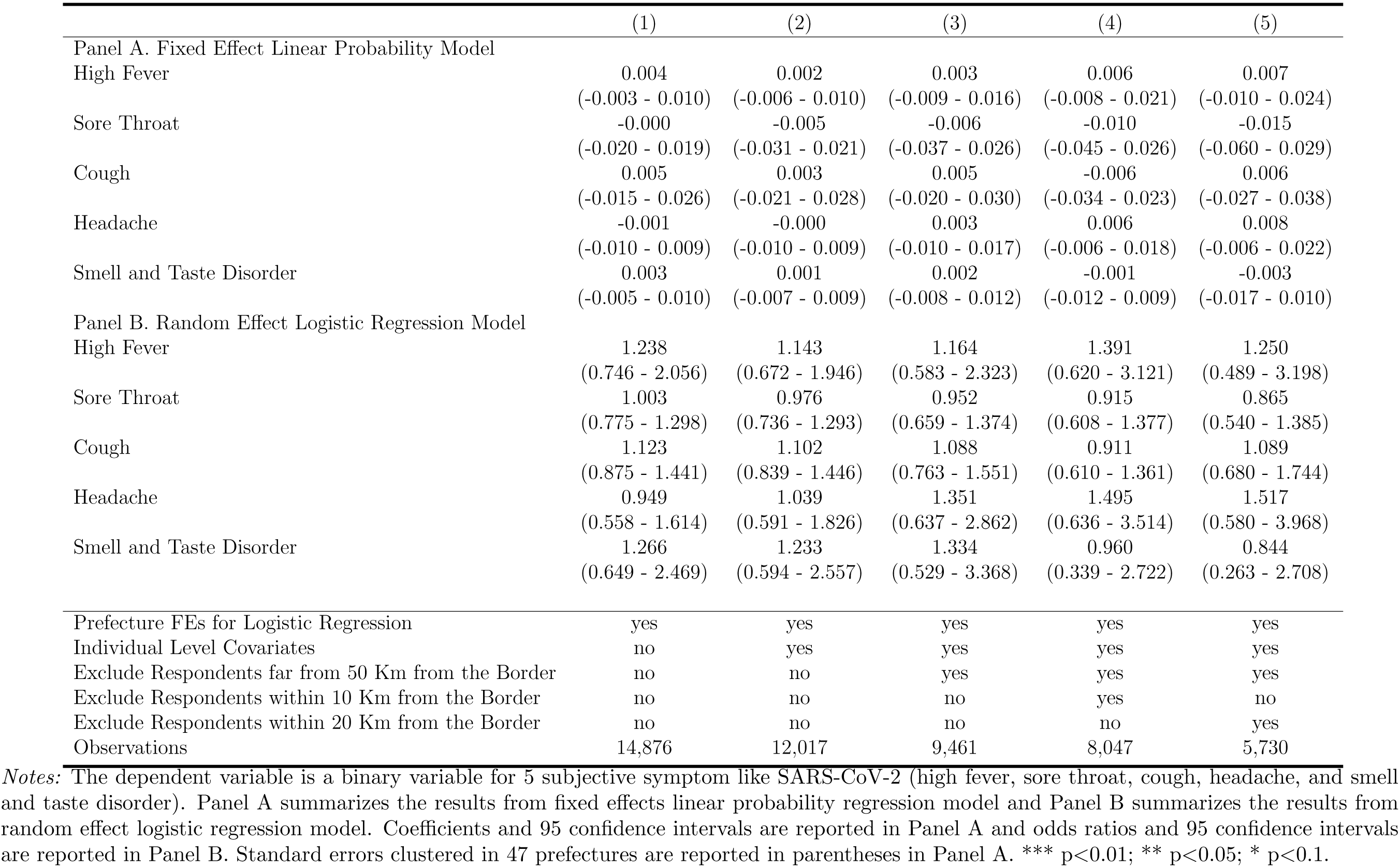
Effects on Symptoms Like SARS-CoV-2 among ”Not Scared”

**Table SI17:**
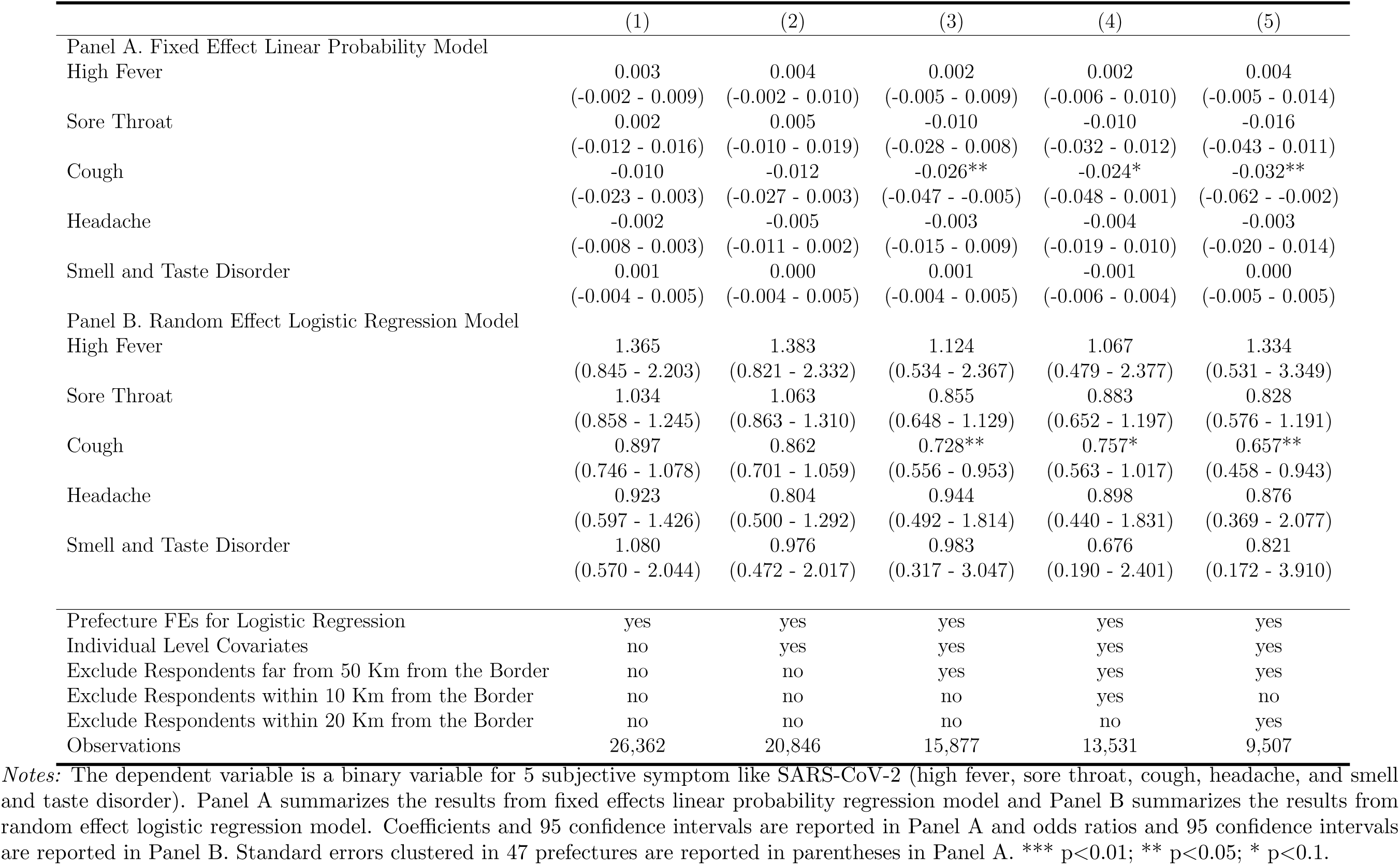
Effects on Symptoms Like SARS-CoV-2 among “Scared”

**Figure SI2:**
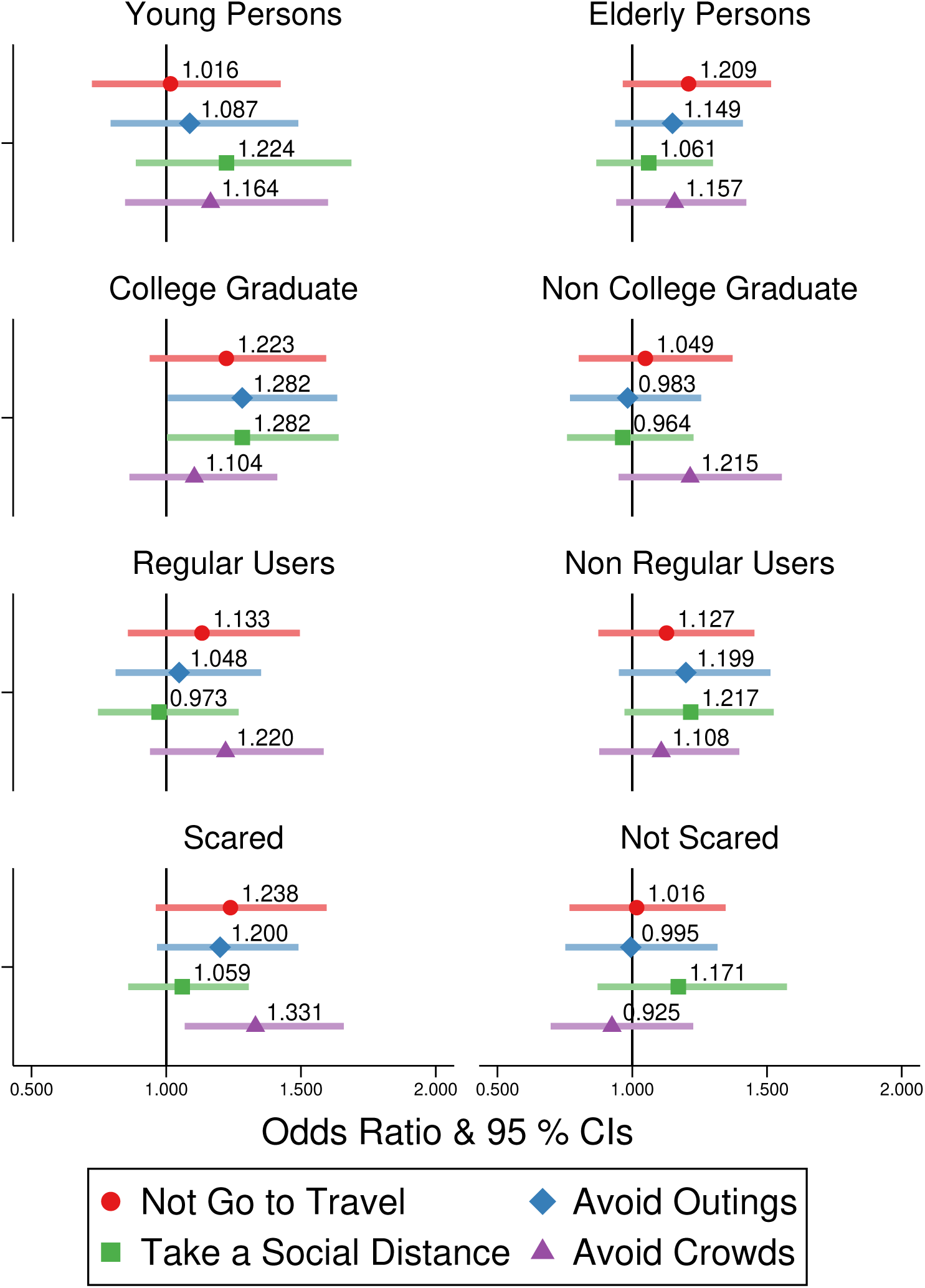
Effects on Going Out Behaviors. *Note:* “Young persons” represents the individuals aged 40 years old or less. “Regular Users” represents the individuals who used Japanese pubs and bars before the pandemic. “Scared” and “Not scared” represent the individuals who reported that they were scared (not scared) at SARS-CoV-2 in the first wave.

